# Development of the FitBack online platform: enhancing global child fitness assessment, health-related interpretation, and surveillance

**DOI:** 10.1101/2024.08.15.24311927

**Authors:** Maroje Sorić, Shawnda A. Morrison, Francisco B. Ortega, Attilio Carraro, Tamás Csányi, Bojan Leskošek, Jarek Mäestu, Snežana Radisavljević-Janić, Luís B. Sardinha, Claude Scheuer, Gregor Starc, Tuija H. Tammelin, Gregor Jurak

## Abstract

**Introduction:** Physical fitness is an important indicator of health in children. Accurately monitoring child and adolescent physical fitness is critical when designing effective public health interventions and strategies. This article aims to provide an overview of the FitBack platform and the health-related criteria that support the reporting system.

**Methods:** Four methodological phases were used to create the FitBack platform. First, a review of existing European fitness databases was conducted to identify common fitness tests and methodologies within a European framework. Second, investigators collated data to create European-level fitness norms, health-related criteria, and health-oriented educational resources dedicated to improving child fitness levels. Third, guidance on how to create one’s own fitness monitoring and surveillance system was created based on good practice examples. The FitBack platform was then piloted and launched based on data received from stakeholders who responded to ≥2 rounds of consultation dialogues.

**Results:** The final version of online platform, with special focus on newly developed Fitback health-related fitness criteria, are communicated herein.

**Discussion:** The FitBack web platform is a first-step towards creating a standardised, person-centred, multi-national fitness surveillance system for children, bridging existing gaps between European political recommendations, individual member states, and harmonious cooperation between sectors. This article also provided the detailed age- and sex-specific health-related fitness cut-points that are used by the FitBack platform, and that could be useful for researchers, practitioners as well as fitness monitoring systems.

## INTRODUCTION

Physical fitness is defined as having the ability to carry out daily tasks with vigour and alertness without undue fatigue, to enjoy leisure time pursuits, and to meet physical challenges inherent in unforeseen emergencies.^1^ Understandably, there is no single measure of physical fitness, but there are different segmentations, e.g., health-related or performance-related fitness. Generally, health-related physical fitness includes cardiorespiratory, musculoskeletal, morphological, and motor components.^2^ Low physical fitness is linked to premature mortality and is a powerful modifiable risk factor to many non-communicable diseases.^3,4^ Moreover, as classical risk factors for non-communicable chronic disease are not typically present before adulthood, physical fitness remains one of the best early indicators of cardiovascular and metabolic health observable in childhood.^5^ Apart from physical health, physical fitness in childhood and adolescence has been consistently associated with higher educational achievement^6^, cognitive abilities, mental health^7^, and lower mortality and disability later in life.^2,8^ Based on the overwhelming evidence higher fitness has on positive health-related outcomes, current European policies encourage member-states to regularly monitor and promote physical fitness in school-aged children.

‘Fitness surveillance refers to the ongoing, systematic, data-collection, analysis, and dissemination of fitness and associated health risk information on a country/regional/municipality level to support public health policy.^9^ So, monitoring child and adolescent physical fitness is critical to determine when creating personalised health strategies^10^, and should not be viewed in any negative context^11^; surveillance is vital for population-based assessment, serving as an early warning system during public health emergencies.^12^ It can document the impact of public health policies, track progress towards specific goals, or observe population trends over time.^13^ Recently, a non-profit network of 10 European partners with expertise in school-based population-level fitness testing was funded by the Erasmus Sport+ scheme (Fitback: The European Network for the Support of Development of Systems for Monitoring Physical fitness of Children and Adolescents). The FitBack project (abb. FitBack), builds on a previous EU-funded project, which validated a safe, reliable and feasible (simple) health-related physical fitness test battery for school-aged children (ALPHA).^14^ FitBack aimed to support fitness surveillance systems at the local, regional or national level, to give individual persons consistent (e.g. annual), timely, and specific health-related feedback on their physical and motor development, in addition to assist in breaking the negative stigma surrounding physical fitness testing in schools.^11^ National-level paediatric physical fitness monitoring systems can better gauge the health status of their society’s population since rapid social changes (e.g., political, technological, environmental) are proving to adversely affect child physical activity.^15–17^ To achieve resilient fitness monitoring and surveillance, several conditions need to be met, including providing fast, accurate, informative health-related feedback to participants to ignite their responsiveness, interest, and continued involvement.

The aim of this article is to provide an overview of the FitBack multi-lingual, web-based, free-for-use platform, which provides personalised, health-related physical fitness feedback for children and adolescents. In addition, in this article, we provide a detail description and ready-to-use age- and sex-specific Fitback health-related fitness criteria that support the FitBack platform and reporting system.

## METHODS

### Literature review of physical fitness testing databases or datasets in Europe

FitBack members performed a thorough review of physical fitness testing systems for children and adolescents and physical fitness-related studies across Europe (e.g., population-based data on physical fitness from research and/or national, regional, or local physical fitness monitoring systems), with the aim to identify suitable datasets that will then be used to update centile values for selected physical fitness tests (stratified by sex and age). The search process was conducted from January – June 2020. We searched for datasets which included the fitness tests outlined in the ALPHA test battery since they are evidence-based, valid, reliable, feasible, safe, and related to health outcomes for children and adolescents.^14^ Specifically, studies conducted between 2000 and 2020 on European children that measured specific components of physical fitness via tests included in ALPHA test battery were included in the search. In addition, partner countries were responsible for performing searches within their own countries by indicating all potential data, either being collected for scientific, or some other purpose, and used at the national level. Searches were performed using PubMed, Scopus, or Google databases. Study parameters extracted included: year of the dataset, age of the subjects (males/females), longitudinal or cross-sectional, data representativeness, fitness test battery, specific fitness tests, raw data availability, country, dataset/study name, organization, contact person, e-mail, and reference to the study.

### FitBack data compilation for creating the underlying fitness database

The final set of data used to comprise the FitBack database included 1,385,174 unique data-points from 109 studies/datasets performed in 36 different countries in European youth aged 6-19 years.^18^ This was supplemented with data previously collected by Tomkinson et al.^19^ These data were included in a FitBack dataset with Monte Carlo simulation used to produce pseudodata (from reported means and SDs) ALPHA tests included: the 20-m shuttle run (cardiorespiratory fitness), handgrip (strength) and standing long jump (muscular power), and body height, body mass, body mass index, and waist circumference (anthropometry). These datasets were then used to produce updated European reference values, European fitness maps, and country rankings. In total, 7,966,693 data points from 34 countries (106 datasets) were made available to create reference values for European children and adolescents aged 6 to 18 years. Weighted averages were calculated across countries and used to create an interactive fitness map of Europe. This work has been described in detail elsewhere.^18^

### Development of health-related criteria for physical fitness tests

To identify optimal available benchmarks for health-related criteria for each item in the ALPHA test battery, we first organised several rounds of discussion during focus groups meetings of partners involved in this stream of work via online platforms. During these discussions, we reached consensus on the main structure and drew the skeleton of the health risk criteria. To identify cut-off values of specific fitness tests that best discriminate children at higher risk of detrimental effects on health, we followed a multi-step procedure. In the first step, we gathered data from partners in the consortium on meta-analytical, or large international studies they have done in this area of research, as well as data on how this is reported in existing fitness surveillance systems. In the next step, for tests that were not covered by the results of stage 1, we performed a scoping literature review. We searched PubMed for studies linking physical fitness in childhood to a health and examining specific values of physical fitness that could best link to poor health outcomes. We applied a harmonised search strategy structured around three areas (1. age group; 2. individual fitness test: waist circumference, 20-m shuttle run, handgrip strength, standing long jump; 3. criterion-based standards and related terms) and limited the search to studies published from 01.01.1980 to 01.07.2020.

We performed the searches by applying a filter for meta-analyses. In the case that no relevant studies were identified in this step, we repeated the search without the filter in the attempt to appropriate individual studies that can be used to identify health risk criteria. We applied a pre-defined list of specific inclusion and exclusion criteria. Data from all included studies were extracted into a custom-made Excel spreadsheet. Multiple studies for specific tests were compared and the best available evidence was selected based on wide age range coverage, and the similarity of the sample to the European population.

Due to space constraints, a more detailed description of the methodology including the extrapolation to missing age periods is presented in the Supplementary materials (Appendix 1).

### Creation of graphical and textual feedback for youth

The first draft of graphical and textual feedback for users was designed based on discussions held at the focus groups involving the FitBack working group, and the feedback received during monthly consortium board meetings. A combination of feedback styles used in the existing national physical fitness monitoring systems (e.g. SLOfit, Netfit, FITescoula, Move!) was used as a starting point for the initial prototypes. This was an iterative process with several prototypes that were being discussed and improved in accordance with the comments received from the working group and the Consortium board. Upon reaching consensus on all features of the graphical and textual feedback, the final prototype was coded and integrated in the web platform. The final prototype was later piloted in six study sites with adolescents. Feedback from end-users was summarised and all suggestions for improvement were reviewed by the working group for inclusion into the final version. To supplement this feedback, we also created easy-to-understand tips for improving each fitness component represented by the ALPHA test battery. First, a template for harmonised structure of tips for improvement of physical fitness was prepared. All comments were collated and discussed during focus groups involving the working group. First drafts of the improvement tips for individual physical fitness components were checked for misalignments in their structure and amended if needed. The final prototypes were coded and integrated into the FitBack web platform.

### Development of an easy-to-use, 10-step guide for establishing fitness surveillance systems

A 10-step guideline on how to design one’s own physical fitness monitoring / surveillance system was created in collaboration with representatives from Finland, Hungary, Portugal, Serbia, and Slovenia – i.e., all countries which have unique experiences in establishing and operating their own national-level physical fitness surveillance and monitoring systems for children. Briefly, after identifying the most robust fitness systems across Europe, the FitBack team conducted numerous rounds of dialogues where common themes emerged, and a detailed SWOT analysis was completed on clustered themes. The final version of the process resulted in these 10-steps: 1) Set up mission statements and aims, 2) Involve stakeholders, 3) Utilize scientific background, 4) Governance structure, 5) Ensure sufficient funding, 6) Data management planning, 7) Provide meaningful feedback, 8) Conduct pilot testing, 9) Plan implementation process, 10) Invest in communication with stakeholders. Details of this process and its outcomes are available in pre-print format.^20^

### FitBack web platform usability and feedback refinement

To confirm the usability of the FitBack web platform we organised several communication activities in all partner countries. Two webpages of the platform were assessed subjectively and objectively in terms of end users’ experiences and usability (i.e., content, navigation, design, and effectiveness) in two phases. Details of the piloting process and the outcomes of the usability evaluation have been described in detail elsewhere.^21^ Briefly, the web page “Make interactive report” was assessed by students and PE teachers, and the webpage “10 steps to design a physical fitness monitoring system” was assessed by policy makers. A total of 111 people from six countries participating in the FitBack project (Croatia, Estonia, Italy, Serbia, Slovenia, and Spain), 77 students, 26 PE teachers, and 13 policy makers provided responses to detailed questionnaires.

### Role of the funding source

The study sponsors had no role in the design and conduct of the study, the analysis and interpretation of data, or in the preparation, review, or approval of the manuscript.

## RESULTS

### Overview of the FitBack web platform

The FitBack web platform includes sections for making interactive fitness reports, resources to improve physical fitness, a section on steps to designing one’s own fitness surveillance system with lessons learned from best-practice established fitness systems, an interactive European fitness map, and an interesting Stories section, which raises awareness on the importance of physical fitness to portal users (Figure 1). The FitBack online platform has been designed to provide educational resources (e.g., infographics on how to improve physical fitness in association with a physical literacy context) to make it a powerful multilingual public health tool, available in the most popular European Union languages: English, Estonian, French, German, Italian and Spanish. Below, we outline the final structure of the platform with a special focus on a key element, the newly developed health-related fitness criteria for a wide age range using scientific literature and interpolations based on the FitBack reference curves.

**Figure 1.** **This Figure contains a photograph of real people and can not be displayed here. The Figure is available at request from the corresponding author (Also available at** https://www.fitbackeurope.eu/en-us/) Fitback European feedback website portal homepage. Reading from left to right, there are drop down menus for “Making a Report”, “Monitoring fitness”, “Fitness Map”, “Stories”, “About” and “Conference”. The top of the FitBack home page depicts the menu structure, language selection and links to one of the basic functions (e.g., interactive report).

### Health-related physical fitness feedback

The main purpose of FitBack was to support the establishment of person-centred physical fitness monitoring systems through the development of a single, multilingual, free, online platform (www.fitbackeurope.eu) that can provide specific, meaningful feedback on individual physical fitness test outcomes which directly relate to health-related criteria which are illustrated in Figure 2.

**Figure 2.**
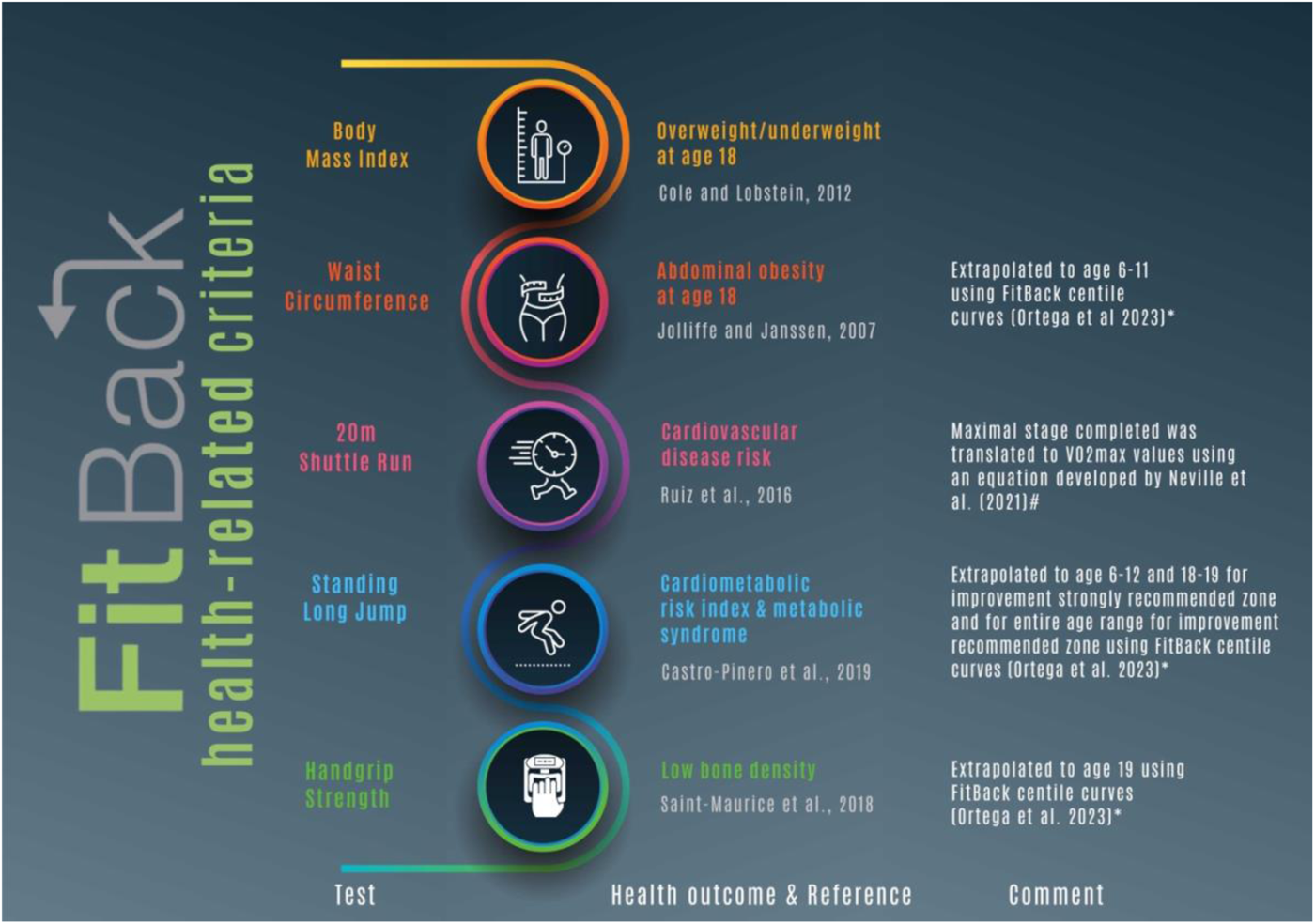
A description of the Fitback health-related criteria for each ALPHA battery test, related health-outcome and scientific evidence-base, and possible additional extrapolation and calculation procedures used.

The first exploration tab the “Make interactive report” so it is easy for users to find and explore. There are two options for entering data into FitBack report module. You can enter data manually into an interactive form that immediately presents a report, or you can import the data for a group of up to 40 students into an Excel template document and generate individual PDF reports. The interactive report is set up so that results of each fitness test are shown in detail along a sliding scale corresponding to the value of the test, i.e., the data entry form (Figure 3) and delivered to the user through a bespoke test evaluation form (Figure 4).

**Figure 3.**
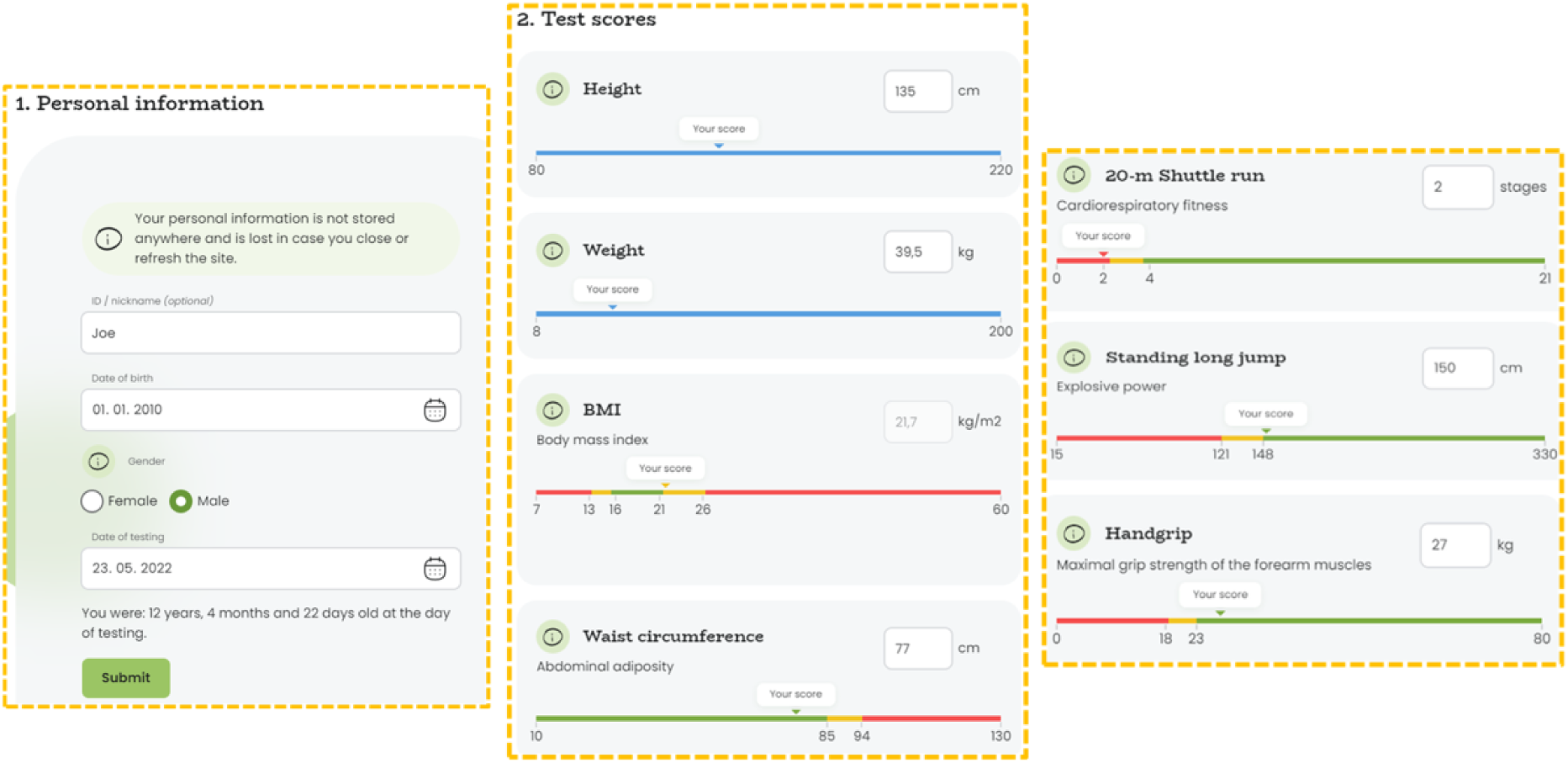
Individual online data entry form for FitBack interactive report. Example of the interactive report information, including the personal data entered to help calculate test score results and relate it to the child’s health risk status. The pane on the left seeks some demographic data (gender and age), and is followed by a screen with entry fields for six ALPHA fit test items (height, weight, waist circumference, 20-m Shuttle run, Standing long jump and Handgrip strength). Note: BMI is automatically calculated from height and weight. *Note: Data on this Figure is completely fictional, created by the authors for illustration purposes only*

**Figure 4.**
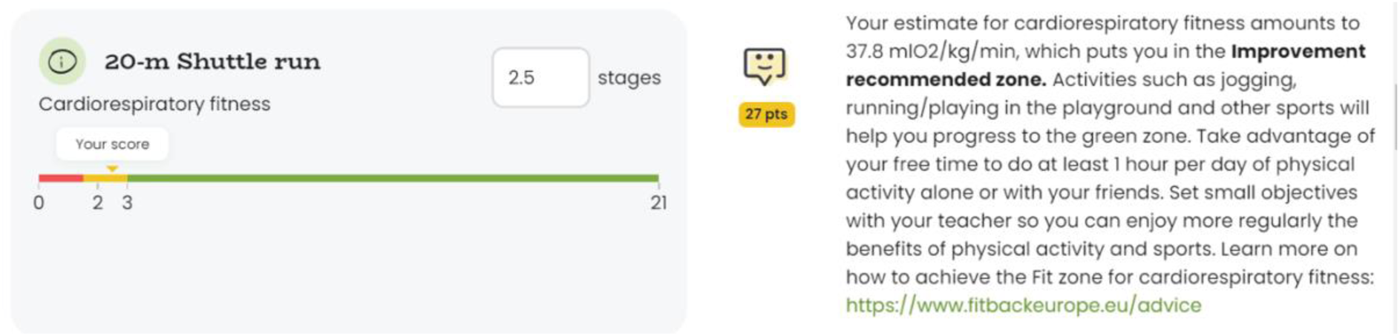
Example of evaluation of test score in interactive report (10 years old boy): as soon as user enters raw score (2.5 stages), health-related (improvement recommended, yellow smiley face) and normative (centile rank, 27 points) evaluation is presented together with text explanation/advice specific for the given fitness level that contains a link to additional info for improving the specific component of physical fitness. The green zone means that the child’s data is within a healthy range. The yellow zone means that some improvements should be made to this item. The red zone means significant improvement is necessary to reach optimal health and well- being. More information is given within-text so the person can follow-up on information should they be interested in learning how they can improve this health metric. *Note: Data on this Figure is completely fictional, created by the authors for illustration purposes only*

Health-related assessments are given based on the best available scientific evidence for that test (Figure 2). The full description of the identified evidence base per specific test is available in the Supplementary material (Table S4), alongside a complete set of “FitBack health-related criteria” values (see Appendix 2; Tables S6-S10). Health-related cut-off points are gender and age specific and are given in a ready-made format to enable convenient future use by academic, education and clinical community. A green zone indicates that the child’s data is within a healthy range whereas a yellow zone means that some improvements should be made to this item. The red zone means significant improvement is necessary to ensure optimal health and well-being. We decided to use “improvement” terminology rather than “health risk zones” or similar terms, since we aimed that the FitBack report is used in a constructive and positive psychological way.

If the user would like more information about a given test, they can click on the information button on the left-hand side of the report, next to the name of the test. A short description will pop up to give the user some additional information on what the test is, and what it is measuring. In the middle of the page, health-related categories will display an emoji face icon to illustrate in which zone the child’s results are located. Points below the emoji denote the percentile rank within EU population (e.g., 45 points indicate that the child has scored higher in this test than 45% of children of the same sex and age across EU). On the right-hand side of the report, detailed information of the implications for health adapted to a given health related zone are described. For even more feedback, parents or interested persons can follow the advice of their PE teacher or share results with the child’s doctor/medical professional.

These interactive reports provide valuable information on the physical fitness level of the child or adolescent which can become an invaluable resource for PE teachers to plan, devise and execute their teaching plan in a more effective and joyful manner. If there are systematic weaknesses in a certain area for a given class of pupils (e.g., poor general fitness, increased body mass index values), FitBack can provide the teacher and student tips for improving specific physical fitness components (Example: www.fitbackeurope.eu/en-gb/create-a-report/-how-can-I-improve-body-composition/) which can increase their physical literacy. Additionally, these recommendations are easy to follow and can be used by adolescents themselves to improve goal-setting, and specific components of their fitness. An example is provided in Figure 5 which includes tips for improving one’s cardiorespiratory fitness.

**Figure 5.**
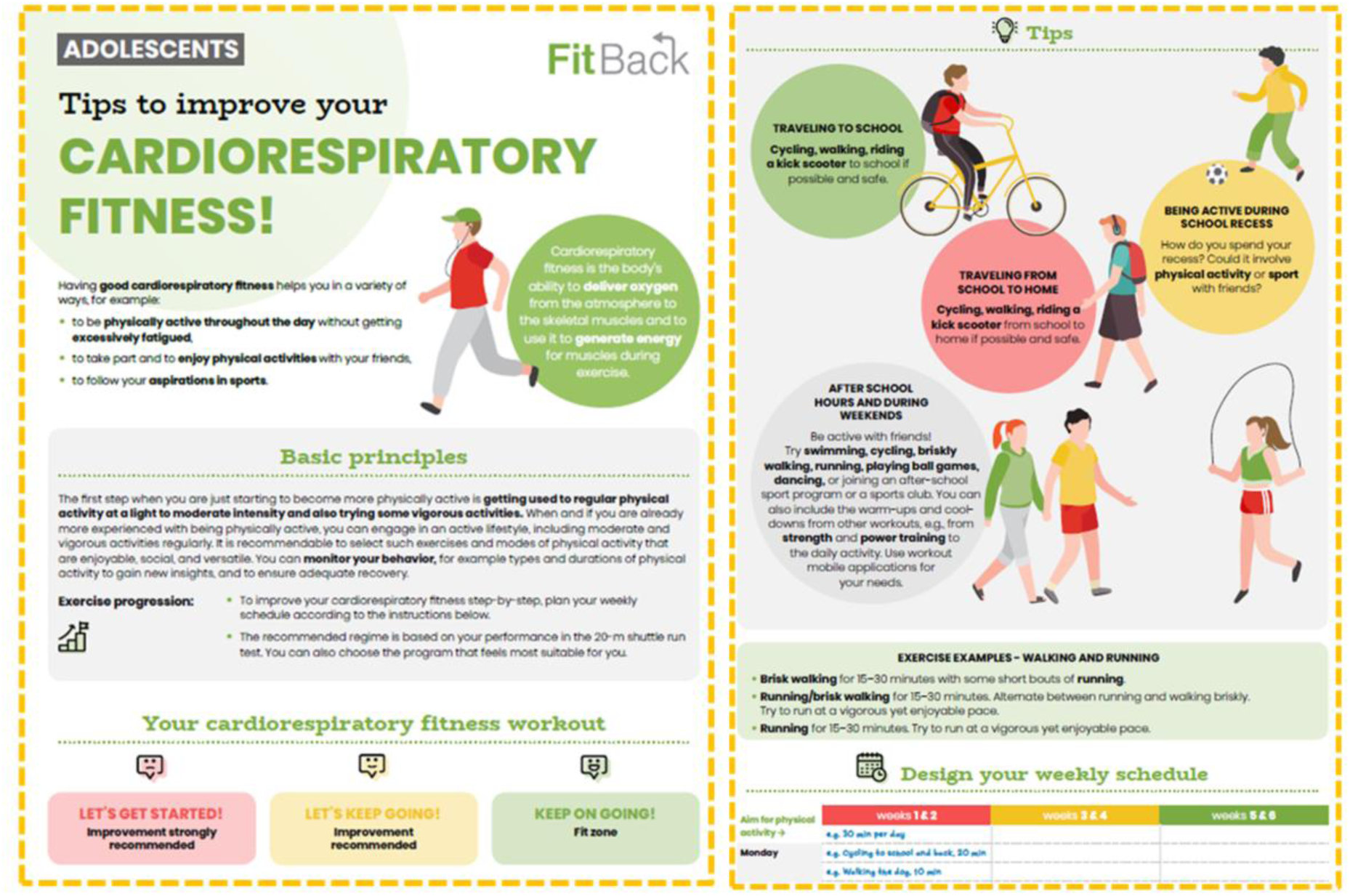
Example of a ‘Tips to improve cardiorespiratory fitness’ document available for download from the FitBack web portal. These support documents are available in multiple language options free of charge to support end users on improving specific physical fitness components.

### Norm-referenced physical fitness feedback

By applying these pre-defined set of inclusion criteria, we retrieved different studies or datasets that included at least one ALPHA fitness test data within the certain age range involving either male, female or both. As previously mentioned, these datasets were used to produce updated European reference values that are used in the fitness reports to provide norm-referenced assessment. In the interactive test report module, alongside health-related evaluation, individual physical fitness test values are also compared to the FitBack-derived European norms^18^, and presented as points on a 0-100 scale, that correspond to a centile value for a given age and gender (Figure 4). In addition, the FitBack web platform provides European fitness maps and country rankings based on the FitBack-derived European norms that can be of interest from a policy making point of view.^18^

### Fitness monitoring or suveillance systems in Europe and steps to design a new one

Another important section in the FitBack web platform is about the fitness monitoring or surveillance systems identified in Europe as good examples of well-established fitness systems, and detailed descriptions of each system have since been published online (https://www.fitbackeurope.eu/en-us/monitoring-fitness/best-practice). In addition, a step by step guide on how to design and establish a national school-based physical fitness monitoring and surveillance system for children and adolescents has been developed and is currently available in a pre-print format.^20^

## DISCUSSION

The FitBack platform physical fitness can provide health-related feedback within individualised fitness reports, supported by other educational materials, for the successful implementation of scalable, person-centred fitness monitoring which can contribute to improving one’s health literacy. In addition, the platform includes descriptions of best practice in fitness monitoring in Europe, and an interactive European fitness map based on up-to-date fitness values for more accurate, evidence-based comparisons between countries and regions. It differs from common national and trans-national surveillance systems (e.g., COSI, etc.) by being person-centred.

### Importance of maintaining adequate childhood fitness for overall health outcomes

Examples of the predictive value of different components of physical fitness to future health have been interpolated by researchers from the FitBack project and others, who have found that both cardiorespiratory fitness and muscular fitness in youth can predict future (⁓25 to 45 years later) chronic and severe disease states^3,8,22,23^ and social and economic consequences related to disability.^24,25^ Higher childhood and adolescent physical fitness is associated with healthier cardiovascular and metabolic profiles, and lower risk of developing^8,26^ CVD later in life. Moreover, improving one’s physical fitness positively affects mental health, and improves overall quality of life.^7^ Recent studies have consistently found that higher fitness is associated with a healthier brain, including structural differences in more grey matter volume in several key regions related to cognition and academic performance^27^, and greater cortical thickness in fitter compared to less fit children.^28^ Likewise, better fitness is associated with better executive function and academic performance.^29,30^ Here, we present the FitBack health-related criteria for fitness assessment across the whole school period and made available the whole set of exact cut-off values for five core fitness tests in the ALPHA-fit test battery, for convenient future use by researchers and clinicians. Currently, the only comparably comprehensive set of health-related criteria is the Fitnessgram system^31^, designed in the U.S. and based on local data. Figures S2 and S3 in Supplementary material show comparison of FitBack and Fitnessgram systems for the two shared fitness tests (i.e., BMI and 20-m Shuttle run). The range of healthy values for BMI are somewhat wider in FitBack standards than in Fitnessgram, especially for age 10-15 yrs. For 20-m shuttle run, the values denoting healthy cardiorespiratory fitness are quite similar in girls, whereas in boys, healthy levels are set higher in FitBack, particularly from ages 10-14.

### On removing barriers to allow for more physical fitness assessments in children

Fitback is a versatile, free-for-use tool which can provide immediate feedback to children, adolescents, parents, teachers, coaches, and health care professionals. This can be used in educational settings for individual or group/class assessments in any European region. Health-related criteria are intended to guide the teaching process by improving population-level health and fitness monitoring through personalised interventions in school settings. It also enables professionals ‘less skilled’ or confident in their fitness assessment and evaluation methods to receive easy-to-understand evidence-based evaluations that can help differentiate risk stratifications between children. By connecting children and their parents to diverse support systems (e.g., teachers, coaches, doctors), FitBack encourages the creation of a unique interdisciplinary ecosystem. FitBack promises to overcome the existing gap between stakeholders who are often working in silos, to encourage a multi-sectorial approach to children’s health and development.

### FitBack Network Support

FitBack is set-up as an open community, welcoming new members interested in advocating for physical fitness as an important biomarker of health in the young. Membership is free and includes: support to countries/regions to establish their own fitness monitoring or surveillance systems, assistance in making the FitBack platform available in other languages, consultations with physical fitness experts in assessment and monitoring, presentations to national key stakeholders and end-users, invitations to special events organised by the FitBack network, and greater networking possibilities (e.g., Active Healthy Kids Global Alliance among others). The FitBack network welcomes researchers who wish to contribute to using or supplying large-scale population data on physical fitness in children.

### Future Directions

Fitness testing amongst school-aged children is often associated with negative experiences and stigma. To this end, The FitBack network is currently leading a follow-up project funded by the EU Erasmus+ Sport Programme called ‘FitBack4Literacy’ which aims to address these negative barriers and provide practical tools for teachers, students, and parents so that everyone will better understand how systematic fitness monitoring can be done in a positive climate and used to support lifelong physical activity habits and enhance health. This project will also enable adding 12 European languages to the existing FitBack web portal content and structure, including languages from migrant and refugee communities to further “promote inclusiveness and diversity, and remove barriers to expert fitness advice”.

Next, and in-line with current EU policies, FitBack will advocate for a ‘life-course’ approach when discussing physical fitness, thereby extending FitBack’s tools and networks to encompass adult and elderly populations. A pilot project is already underway in Slovenia^32^, after which results will be scaled-up to the European level.

## Conclusion

Current scientific evidence supports the importance of evaluating physical fitness to enhance public health, education, and sport performance. The FitBack platform allows children, parents, researchers, PE teachers, physicians, policymakers, and others to accurately interpret physical fitness levels of children and adolescents in a more accessible way. This article also provided a set of the most current, robust, and evidence-based sex and age specific health-related criteria for five ALPHA fit tests adopted by FitBack. Health-related criteria are given in a ready-made format that can be useful for researchers, health and sport practitioners, as well as implemented in current and future regional or nation-wide fitness monitoring and surveillance systems. Countries can translate the platform into their own language(s), they can join the FitBack network to receive mentorship, guidance, and feedback from experts. The FitBack legacy aims to continue promoting physical fitness and physical literacy across the lifespan to meet the demands of the multiple, dynamic global challenges existing today.

## FUNDING

This research has been co-financed by: the Erasmus+ Sport Program of the European Union within the framework of the FitBack project (n° 613010-EPP-1-2019-1-SI-SPO-SCP), the Slovenian Research and Innovation Agency (Bio-psycho-social research program, n° P5-0142) and the Scientific Unit of Excellence in Exercise, Nutrition and Health (UCEENS) funded by the University of Granada’s Own Research Plan. The study sponsors had no role in the design and conduct of the study, the analysis and interpretation of data, or in the preparation, review, or approval of the manuscript.

## CONFLICTS OF INTEREST

The authors declare no conflicts of interest for the present work.

## AUTHORSHIP CONTRIBUTIONS

Conceptualization: MS, SAM, FBO, GJ; Writing - Original Draft: SM, MS, GJ; Writing - Review & Editing: MS, SAM, FBO, AC, TC, BL, JM, SRJ, LBS, CS, GS, TT, GJ; Methodology: MS; Visualizations: SAM, BL, GJ; Project administration: GJ; Funding acquisition: GJ; Supervision: MS, GJ

## DATA AVAILABILITY STATEMENT

Data supporting the findings of this study are available within the the supplementary materials.

## ETHICAL APPROVAL

Not applicable

## Notes

### Competing Interest Statement

The authors have declared no competing interest.

