## Supplementary materials for "Development of the FitBack online platform: enhancing global child fitness assessment, health-related interpretation, and surveillance"

### 1. Development of the FitBack health-related criteria for each of the components of physical fitness

To identify cut-off values of specific fitness tests that best discriminate children under higher risk of detrimental effects on health we followed a multi-step procedure. In the first step we gathered data from partners in the consortium on meta-analytical, or large international studies they have done in this area of research, as well as data on how this is reported in existing fitness surveillance systems. After reaching out to every individual partner in the consortium to collect systematic reviews or meta-analyses that have examined relations of fitness, we identified one such study performed within the consortium, and related to 20-m Shuttle run test (Ruiz et al, 2016). In the next step we gathered data from partners in the consortium on how this health-related fitness zones are reported in existing fitness surveillance systems. The results of this exercise are summarised in Table S1. Criterion-referenced evaluation is available in at least one of the existing surveillance systems for four fitness tests (BMI, waist circumference, 20-m Shuttle run, Handgrip strength), but not for Standing long jump. It should be noted that, while criterion-referenced standards for body composition and abdominal obesity (i.e., BMI and waist circumference, respectively) are based on a large international, or nationally representative studies, criteria for other two tests (20-m Shuttle run, Handgrip strength) come from small-to-medium-scale individual studies. Hence, of all the sources identified during this process, only BMI criteria were deemed suitable for direct inclusion in the FitBack platform. Since two different sets of criteria standards for BMI were identified across existing surveillance systems, a discussion during a Consortium board meeting was scheduled. Based on the results of this discussion, IOTF criteria for BMI were chosen to be included in the FitBack health-related fitness standards (Cole and Lobstein, 2012).

**Table S1. Reference values for health-related fitness components used in existing fitness surveillance systems**

|  | **SLOfit** | **FITescola** | **NETFIT** | **MOVE!** | **Serbian fitness surveillance** |
| --- | --- | --- | --- | --- | --- |
| **BMI** | CR (IOTF) | CR (WHO) | CR (IOTF) | / | / |
| **Waist circumference** | / | CR (Age-Specific Adolescent Metabolic Syndrome Criteria) | / | / | / |
| **20-m Shuttle run** | / | CR standards (Fitnessgram) | CR standards (Fitnessgram) | NR (tertiles) | NR (centiles) based on national data |
| **Handgrip strength** | / | / | CR standards (Iowa Bone Development Study) | / | / |
| **Standing long jump** | NR (centiles) based on national data | NR (centiles) based on international data | NR (centiles) based on national data | / | NR (centiles) based on national data |

*CR=criterion-referenced; NR= norm-referenced*

In the next step, for tests that were not covered by the results of stage 1, we performed a scoping literature review. We searched Pubmed for studies linking physical fitness in childhood to a health outcome, and examining specific values of physical fitness that can best identify poor health outcomes. We applied a harmonised search strategy structured around 3 areas (1. age group; 2. individual fitness test: waist circumference, 20-m shuttle run, handgrip strength, standing long jump; 3. criterion-based standards and related terms) and limited the search to studies published from 1980 to 01.07.2020. We first performed the searches by applying a filter for meta-analyses. In case no relevant studies were identified in this step, we repeated the search without the filter in the attempt to appropriate individual studies that can be used to identify health risk criteria. Specific inclusion and exclusion criteria are given in table S2, while a PRISMA flowchart describing this process in more detail for standing long jump is presented in Figure S1 as an example of the full search process.

**Table S2. Inclusion and exclusion criteria set for the search strategy.**

| **Inclusion criteria:** | 1. study investigated the ability of a specific test as predictor of metabolic, cardiovascular or bone health.  2. study reports data of healthy children and adolescents (aged between 6 and 19 years, or part of this age group, or mean age in this interval).  3. one of the tests from the alpha-fit testing battery measured  4. a fitness cut point associated with disease risk was calculated in the study; |
| --- | --- |
| **Exclusion criteria:** | 1. studies with specific groups of patients with obesity or other conditions |

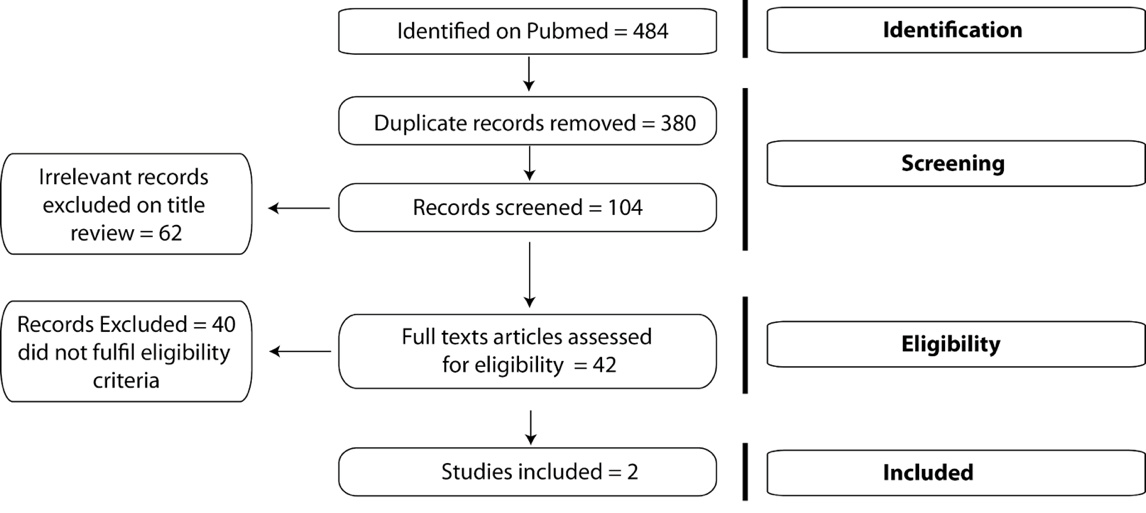

**Figure S1. An example of a study search process showing a PRISMA Flowchart for standing long jump test**

Table S3 summarises the results of the screening process for four tests included in this exercise. Data from all included studies (i.e., 68 altogether) were extracted into a custom made excel spreadsheet. Multiple studies for specific tests were compared and the best available evidence was selected based on wide age range coverage, and the similarity of the sample to the European population.

**Table S3. Summary of the results of the screening process for four tests included in this exercise.**

| **Fitness test** | **No of meta-analysis screened (included)** | **No of individual studies screened (included)** |
| --- | --- | --- |
| **Waist circumference** | 0 | 2197 (59 included) |
| **20-m Shuttle run** | 1 (1 included)* | N.A. |
| **Handgrip strength** | 0 | 880 (6 included) |
| **Standing long jump** | 0 | 104 (2 included) |

**since a meta-analysis published in 2016 was identified in Step 1 for 20-m Shuttle run, we searched only for more recent meta-analyses. However, no additional meta-analyses from 2016 to July 2020 were identified; N.A.: not applicable.*

Optimal sources for criterion-based, health-related fitness standards identified are shown in Table S4, along with related study characteristics. Most of these studies link physical fitness with cardiovascular or metabolic health. A notable exception is the study by Castro-Pinero et al. (2019) that associated leg power (measured by standing long jump) to bone health.

**Table S4. Studies chosen for criterion-based FitBack health-related fitness standards and their basic characteristics.**

| **Fitness test** | **Reference** | **Design** | **Population** | **Fitness measurement** | **Health outcome** | **Data analysis** | **Metrics** |
| --- | --- | --- | --- | --- | --- | --- | --- |
| BMI | Cole and Lobstein, 2012 | 6 Longitudinal studies | 192 727 children (97 876 males) from six countries (UK, USA, the Netherlands, Brazil, Singapore and Hong Kong) aged between 2 and 18 years. | Body composition; measured height and weight | BMI at age 18 | Averaged L, M and S curves (curves from the 6 countries were used to derive new cut-offs defined in terms of the centiles at 18 years corresponding to each BMI value).  These new cut-offs were compared with the originals, and with the WHO standard and  Reference (based on US and Chinese data). | The new cut-offs were virtually identical to the originals, giving prevalence rates differing by <0.2% on average.  The discrepancies were smaller for overweight and obesity than for thinness. |

| **Fitness test** | **Reference** | **Design** | **Population** | **Fitness measurement** | **Health outcome** | **Data analysis** | **Metrics** |
| --- | --- | --- | --- | --- | --- | --- | --- |
| Waist circumference | Jolliffe and Janssen, 2007 | The NHANES nationally representative cross-sectional surveys  (conducted with a stratified multistage probability design of the U.S. population). | 2- to 20-year-old adolescents (2,921 male and 3,146 female participants) | Body composition; waist circumference measured to the nearest 0.1 cm at the level of the iliac crest with standard protocols | Abdominal obesity at age 19 | growth curves analysis | Age- and gender-specific cut-points for each MetS  component were developed that can be used to define high-risk values in 12- to 19-year-olds. |

| **Fitness test** | **Reference** | **Design** | **Population** | **Fitness measurement** | **Health outcome** | **Data analysis** | **Metrics** |
| --- | --- | --- | --- | --- | --- | --- | --- |
| **20-m Shuttle run test** | Ruiz et al., 2016 | Systematic review with meta-analysis  (included studies with no restrictions were considered regarding study design or data collection) | 9280 children and adolescents (4717 boys and 4563 girls) aged 8–19 years from 14 countries | cardiorespiratory fitness was assessed by a submaximal or maximal exercise test and can be expressed as VO2max (mL/kg/min) with different protocols. VO2max was estimated by different formulas (Leger, Hansen, etc.) | Cardiovascular disease risk  Inclusion criteria: 1. cardiorespiratory fitness cut point associated with cardiovascular disease risk was calculated in the study; 2. at least two cardiovascular disease risk factors were measured and 3. definition of cardiovascular disease risk are provided. | area under the curve (AUC) | META-ANALYSIS RESULTS:  The pooled dOR estimates were 5.7 (95% CI 4.8 to 6.7) for boys and 3.6 (95% CI 3.0 to 4.3) for girls.  The area under the HSROC curve estimating the discriminating accuracy of cardiorespiratory fitness for identifying cardiovascular disease risk was 0.71 (95% CI 0.65 to 0.76; p<0.001) in boys and 0.64 (95% CI 0.58 to 0.69; p<0.001) in girls. |

| **Fitness test** | **Reference** | **Design** | **Population** | **Fitness measurement** | **Health outcome** | **Data analysis** | **Metrics** |
| --- | --- | --- | --- | --- | --- | --- | --- |
| **Standing long jump** | Castro-Pinero et al., 2019 | Cross-sectional study | 969 adolescents (aged 12.5–17.5 years old) from 9 European countries (520 girls). | The SLJ was performed from a starting position immediately behind a line, standing with feet approximately shoulder’s width apart, and the adolescent jumped as far forward as possible, landing with their feet together. The test was performed twice, with 1-minute rest between the measurements, and the longest distance achieved was recorded in centimeters. | 1. metabolic syndrome, determined using WC, diastolic and systolic blood pressure, TG, HDL, and fasting glucose with the definition by Jolliffe and Janssen (2007)  2. cardiometabolic risk factors were expressed as age- and sex-specific z-scores A continuous cardiometabolic risk index was computed as the average z-score of four risk factors (WC, mean arterial pressure, TG/HDL ratio, and fasting insulin). The adolescents were categorized as having elevated cardiometabolic risk if their cardiometabolic risk index was one standard deviation above the mean for each of the four markers | Receiver operating characteristic (ROC) analyses  ideal cut points for each of the four tests, were based on the Youden Index, but also based on the PPV, NPV, and diagnostic OR for each threshold. ROC analyses were completed for all four muscle strength parameters: absolute HG, relative HG(HG / body mass), absolute SLJ, and relative SLJ (SLJ × body mass).The Least Mean Square (LMS) method was used to create age-and sex-specific z-scores and were used in the main analysis. | *1) ROC-derived cut points and diagnostic statistics for tests of muscle strength to determine elevated cardiometabolic risk indeks: (YI=* Youden index)  BOYS: Standing long jump (cm): Cut point Z-score: ≤ −0.790; YI: 0.54  GIRLS: Standing long jump (cm): Cut point Z-score: ≤ −0.846; YI: 0.28  *2) ROC-derived cut points and diagnostic statistics for tests of muscle strength to determine metabolic syndrome (YI= Youden index)*  BOYS: Standing long jump: Cut point Z-score: ≤ −0.890; YI: 0.44  GIRLS: Standing long jump (cm): Cut point Z-score: ≤ −0.797; YI: 0.37 |

| **Fitness test** | **Reference** | **Design** | **Population** | **Fitness measurement** | **Health outcome** | **Data analysis** | **Metrics** |
| --- | --- | --- | --- | --- | --- | --- | --- |
| **Handgrip strength** | Saint-Maurice et al., 2018 | Repeated measure cohort study | 228, 241, 200, and 168 males, and 245, 244, 213, and 196 females measured at age 11, 13, 15 and 17, respectively | Grip strength was measured using a standard protocol for the handgrip test (Jamar; Creative Health Products, Plymouth, MI; elbow flexed at 90°). The best score of two trials obtained from the dominant hand was re- corded for analysis. | Low bone density  Total body (less the head) bone mineral content (TBLH_BMC; g) was measured using a standardized protocol for dual- energy x-ray absorptiometry (DXA)  Insufficient TBLH_BMC was defined using two thresholds, more specifically, Z < − 0.84 (i.e., below 20th percentile) and Z < − 1.5 (below ~ 7th percentile) for their age and sex. | Receiver Operator Characteristic (ROC) curves.  Se was emphasized (i.e., ≥ 80.0) for -0.84Z cut-off while preserving Sp to 60.0 or higher.  For -1.5Z cut-off Sp was emphasized (i.e., ≥ 80.0) while preserving Se to 60.0 or higher. | The AUC approximated or reached 0.80 for both cut-offs in both males and females (Males: -0.84Z AUC = 0.71, -1.5Z AUC = 0.80; Females: -0.84Z AUC = 0.81, -1.5Z AUC = 0.79).  Boys: -0.84Z cut-off associated with handgrip Z-score of − 0.10 (Se = 81.4, Sp = 53.6)  -1.5Z cut-off associated with handgrip Z-score of − 1.13 (Se = 80.0, Sp = 82.4).  Girls: -0.84Z cut-off associated with handgrip Z-score of − 0.11 (Se = 92.5, Sp = 54.1)  -1.5Z cut-off associated with handgrip Z-score of − 1.23 (Se = 63.6, Sp = 80.9) |

As it can be deduced from Table S4 several issues with identified sources were acknowledged that prevented direct implementation of identified cut-off values in the FitBack platform:

1. Sources for several tests (i.e., waist circumference, 20-m Shuttle run, Handgrip strength, Standing long jump) do not cover the whole age range included in the FitBack platform.
2. Criteria for standing long jump allow the identification of only 2 health zones, while all other standards classify individuals in 3 distinct groups.
3. Criteria for 20-m Shuttle run are related to VO_2max_, hence translation of maximal speed attained/stage completed to VO_2max_ is necessary.

Ad 1) For missing age groups issue, we used recently developed FitBack centile curves (Ortega et al.,2023) to first identify the centile value of the youngest and/or oldest available age that corresponds to health-related criterium value. After that, we extrapolated it to younger and/or older ages by finding raw test values for the corresponding FitBack percentile for each age and sex group. Specific centile values for individual tests across categories of health-related criteria are shown in Table S5.

**Table S5. Summary of age ranges for which criteria had to be extrapolated and FitBack centile values of the youngest and/or oldest available age that corresponds to health-related criterium value for a given fitness test**

| **Fitness test** | **age ranges extrapolated** | **Improvement strongly recommended zone** | | **Improvement recommended zone** | |
| --- | --- | --- | --- | --- | --- |
|  |  | boys | girls | boys | girls |
| **Waist circumference** | 6-11 | 96. centile | 86. centile | 90. centile | 68. centile |
| **Standing long jump** | 6-12 and  18-19 | 13. centile  24. centile | 10. centile  16. centile | 40. centile*  40. centile* | 40. centile*  40. centile* |
| **Handgrip strength** | 19 | 11. centile | 18. centile | 37. centile | 49. centile |

**values extrapolated for the entire 6-19 age range as described in the main text*

Ad 2) For missing level issue for Standing long jump test, we first detected FitBack centiles that corresponded to criterium values reported in the source identified earlier (Castro-Pinero et al., 2019). As this approximated 10. centile, we looked for studies that have evaluated associations of lower body strength in youth with hard end-points across categories of muscle strength. More specifically, we repeated the search in Pubmed by using previous search string for standing long jump test while replacing criterion-based search area with different wordings for normative evaluation. Through this process, the best available evidence for associations of lower body strength with health was identified (Ortega et al., 2012). This analysis that included male adolescents showed that 10% of youth with the lowest strength are at the highest risk for premature death and that the gradual reduction in risk is seen across 3 next deciles. After the 4^th^ decile no additional benefits were seen. Very similar patterns were observed for the association of strength in adolescence and disability 30 years after (Henrikson H et al., 2019). Based on this, 10. and 40. centile were selected as cut-off points for the “Improvement strongly recommended” and “Improvement recommended” zone, respectively. Correspondingly granular analysis in women was not identified, but existing data indicate that the association between strength and mortality is even stronger in women compared to men (Garcia-Hermoso et al., 2018). Hence, identical FitBack centile values were proposed as health-related criterion standards for girls as for boys.

Ad 3) In order to translate maximal speed attained/stage completed during a 20-m Shuttle run test to VO_2max_ values we searched Pubmed for available equations in children. The search identified 6 different equations. Data from all studies were extracted into a custom made excel spreadsheet, the equations were compared, and the optimal one was selected based on wide age and sex range coverage as well as plausible variability for various stages completed across age groups. Based on these criteria we opted for an equation developed by Neville et al. (2021). This group developed an allometric model based on data from 306 9-18 year old children which explained almost 70% of the variance with the standard error of estimate SEE = 0.11 (or 11.6%). The equation is given below:

*VO_2max_ (ml/kg/min)=M^-0.126^ + S^1.531^ × exp (0.808 − 0.116 × girls), where M = Mass and S = 20 mSRT running speed (km/h) and “girls” is entered as a [0,1] indicator variable (boys = 0 and girls = 1).*

**2. A complete set of cut-off values for Fitback health-related criteria across five ALPHA-fit fitness tests (Body mass index, waist circumference, 20-m Shuttle run, Standing long jump, Handgrip strength)**

Complete set of cut-off values for FitBack health-related criteria across five ALPHA-fit fitness tests is given in Tables S6-S10. All values are stratified by sex and age group.

**Table S6. FitBack health-related reference values for Body mass index (kg/m2) by age group, stratified by sex (Based on Cole, T. J., & Lobstein, T. (2012). Extended international (IOTF) body mass index cut-offs for thinness, overweight and obesity. Pediatric Obesity, 7(4), 284-294. doi:10.1111/j.2047-6310.2012.00064.x)**

| **Age (years)** | **Improvement strongly recommended** | **Improvement recommended** | **Fit Zone** | **Improvement recommended** | **Improvement strongly recommended** |
| --- | --- | --- | --- | --- | --- |
| **GIRLS** | | | | | |
| 5.08 | <12.56 | 12.57-14.02 | 14.03-17.22 | 17.23-19.21 | >19.22 |
| 5.17 | ≤12.54 | 12.55-14 | 14.01-17.22 | 17.23-19.23 | ≥19.24 |
| 5.25 | ≤12.52 | 12.53-13.98 | 13.99-17.22 | 17.23-19.26 | ≥19.27 |
| 5.33 | ≤12.5 | 12.51-13.97 | 13.98-17.23 | 17.24-19.29 | ≥19.3 |
| 5.42 | ≤12.48 | 12.49-13.95 | 13.96-17.23 | 17.24-19.32 | ≥19.33 |
| 5.5 | ≤12.45 | 12.46-13.93 | 13.94-17.24 | 17.25-19.35 | ≥19.36 |
| 5.58 | ≤12.43 | 12.44-13.92 | 13.93-17.25 | 17.26-19.39 | ≥19.4 |
| 5.67 | ≤12.41 | 12.42-13.9 | 13.91-17.26 | 17.27-19.42 | ≥19.43 |
| 5.75 | ≤12.39 | 12.4-13.89 | 13.9-17.27 | 17.28-19.47 | ≥19.48 |
| 5.83 | ≤12.37 | 12.38-13.87 | 13.88-17.29 | 17.3-19.51 | ≥19.52 |
| 5.92 | ≤12.36 | 12.37-13.86 | 13.87-17.3 | 17.31-19.56 | ≥19.57 |
| 6 | ≤12.34 | 12.35-13.85 | 13.86-17.32 | 17.33-19.6 | ≥19.61 |
| 6.08 | ≤12.32 | 12.33-13.84 | 13.85-17.34 | 17.35-19.66 | ≥19.67 |
| 6.17 | ≤12.31 | 12.32-13.83 | 13.84-17.36 | 17.37-19.71 | ≥19.72 |
| 6.25 | ≤12.29 | 12.3-13.82 | 13.83-17.38 | 17.39-19.77 | ≥19.78 |
| 6.33 | ≤12.28 | 12.29-13.82 | 13.83-17.41 | 17.42-19.83 | ≥19.84 |
| 6.42 | ≤12.27 | 12.28-13.81 | 13.82-17.44 | 17.45-19.89 | ≥19.9 |
| 6.5 | ≤12.26 | 12.27-13.81 | 13.82-17.47 | 17.48-19.95 | ≥19.96 |
| 6.58 | ≤12.25 | 12.26-13.81 | 13.82-17.5 | 17.51-20.02 | ≥20.03 |
| 6.67 | ≤12.24 | 12.25-13.81 | 13.82-17.53 | 17.54-20.09 | ≥20.1 |
| 6.75 | ≤12.23 | 12.24-13.81 | 13.82-17.57 | 17.58-20.16 | ≥20.17 |
| 6.83 | ≤12.23 | 12.24-13.81 | 13.82-17.6 | 17.61-20.23 | ≥20.24 |
| 6.92 | ≤12.23 | 12.24-13.82 | 13.83-17.64 | 17.65-20.31 | ≥20.32 |
| 7 | ≤12.23 | 12.24-13.83 | 13.84-17.68 | 17.69-20.38 | ≥20.39 |
| 7.08 | ≤12.23 | 12.24-13.83 | 13.84-17.72 | 17.73-20.46 | ≥20.47 |
| 7.17 | ≤12.23 | 12.24-13.84 | 13.85-17.77 | 17.78-20.54 | ≥20.55 |
| 7.25 | ≤12.23 | 12.24-13.86 | 13.87-17.81 | 17.82-20.62 | ≥20.63 |
| 7.33 | ≤12.24 | 12.25-13.87 | 13.88-17.86 | 17.87-20.71 | ≥20.72 |
| 7.42 | ≤12.24 | 12.25-13.88 | 13.89-17.9 | 17.91-20.79 | ≥20.8 |
| 7.5 | ≤12.25 | 12.26-13.9 | 13.91-17.95 | 17.96-20.88 | ≥20.89 |
| 7.58 | ≤12.25 | 12.26-13.91 | 13.92-18 | 18.01-20.97 | ≥20.98 |
| 7.67 | ≤12.26 | 12.27-13.93 | 13.94-18.06 | 18.07-21.06 | ≥21.07 |
| 7.75 | ≤12.27 | 12.28-13.95 | 13.96-18.11 | 18.12-21.15 | ≥21.16 |
| 7.83 | ≤12.28 | 12.29-13.96 | 13.97-18.16 | 18.17-21.24 | ≥21.25 |
| 7.92 | ≤12.29 | 12.3-13.98 | 13.99-18.22 | 18.23-21.34 | ≥21.35 |
| 8 | ≤12.3 | 12.31-14 | 14.01-18.27 | 18.28-21.43 | ≥21.44 |
| 8.08 | ≤12.31 | 12.32-14.02 | 14.03-18.33 | 18.34-21.53 | ≥21.54 |
| 8.17 | ≤12.32 | 12.33-14.04 | 14.05-18.38 | 18.39-21.63 | ≥21.64 |
| 8.25 | ≤12.33 | 12.34-14.06 | 14.07-18.44 | 18.45-21.73 | ≥21.74 |
| 8.33 | ≤12.34 | 12.35-14.08 | 14.09-18.5 | 18.51-21.83 | ≥21.84 |
| 8.42 | ≤12.35 | 12.36-14.1 | 14.11-18.56 | 18.57-21.93 | ≥21.94 |
| 8.5 | ≤12.37 | 12.38-14.12 | 14.13-18.62 | 18.63-22.03 | ≥22.04 |
| 8.58 | ≤12.38 | 12.39-14.15 | 14.16-18.68 | 18.69-22.13 | ≥22.14 |
| 8.67 | ≤12.39 | 12.4-14.17 | 14.18-18.74 | 18.75-22.23 | ≥22.24 |
| 8.75 | ≤12.4 | 12.41-14.19 | 14.2-18.8 | 18.81-22.34 | ≥22.35 |
| 8.83 | ≤12.41 | 12.42-14.21 | 14.22-18.86 | 18.87-22.44 | ≥22.45 |
| 8.92 | ≤12.42 | 12.43-14.23 | 14.24-18.92 | 18.93-22.55 | ≥22.56 |
| 9 | ≤12.44 | 12.45-14.26 | 14.27-18.98 | 18.99-22.65 | ≥22.66 |
| 9.08 | ≤12.45 | 12.46-14.28 | 14.29-19.04 | 19.05-22.76 | ≥22.77 |
| 9.17 | ≤12.46 | 12.47-14.3 | 14.31-19.11 | 19.12-22.87 | ≥22.88 |
| 9.25 | ≤12.47 | 12.48-14.33 | 14.34-19.17 | 19.18-22.98 | ≥22.99 |
| 9.33 | ≤12.49 | 12.5-14.35 | 14.36-19.23 | 19.24-23.08 | ≥23.09 |
| 9.42 | ≤12.5 | 12.51-14.38 | 14.39-19.3 | 19.31-23.19 | ≥23.2 |
| 9.5 | ≤12.52 | 12.53-14.4 | 14.41-19.37 | 19.38-23.3 | ≥23.31 |
| 9.58 | ≤12.53 | 12.54-14.43 | 14.44-19.43 | 19.44-23.41 | ≥23.42 |
| 9.67 | ≤12.55 | 12.56-14.46 | 14.47-19.5 | 19.51-23.52 | ≥23.53 |
| 9.75 | ≤12.57 | 12.58-14.49 | 14.5-19.57 | 19.58-23.63 | ≥23.64 |
| 9.83 | ≤12.59 | 12.6-14.52 | 14.53-19.63 | 19.64-23.74 | ≥23.75 |
| 9.92 | ≤12.61 | 12.62-14.55 | 14.56-19.7 | 19.71-23.85 | ≥23.86 |
| 10 | ≤12.63 | 12.64-14.58 | 14.59-19.77 | 19.78-23.96 | ≥23.97 |
| 10.08 | ≤12.65 | 12.66-14.61 | 14.62-19.84 | 19.85-24.07 | ≥24.08 |
| 10.17 | ≤12.67 | 12.68-14.64 | 14.65-19.91 | 19.92-24.18 | ≥24.19 |
| 10.25 | ≤12.69 | 12.7-14.68 | 14.69-19.98 | 19.99-24.28 | ≥24.29 |
| 10.33 | ≤12.72 | 12.73-14.71 | 14.72-20.06 | 20.07-24.39 | ≥24.4 |
| 10.42 | ≤12.74 | 12.75-14.75 | 14.76-20.13 | 20.14-24.5 | ≥24.51 |
| 10.5 | ≤12.77 | 12.78-14.78 | 14.79-20.2 | 20.21-24.61 | ≥24.62 |
| 10.58 | ≤12.79 | 12.8-14.82 | 14.83-20.27 | 20.28-24.71 | ≥24.72 |
| 10.67 | ≤12.82 | 12.83-14.86 | 14.87-20.35 | 20.36-24.82 | ≥24.83 |
| 10.75 | ≤12.85 | 12.86-14.9 | 14.91-20.42 | 20.43-24.93 | ≥24.94 |
| 10.83 | ≤12.88 | 12.89-14.94 | 14.95-20.5 | 20.51-25.03 | ≥25.04 |
| 10.92 | ≤12.91 | 12.92-14.98 | 14.99-20.57 | 20.58-25.14 | ≥25.15 |
| 11 | ≤12.94 | 12.95-15.03 | 15.04-20.65 | 20.66-25.24 | ≥25.25 |
| 11.08 | ≤12.97 | 12.98-15.07 | 15.08-20.72 | 20.73-25.35 | ≥25.36 |
| 11.17 | ≤13.01 | 13.02-15.11 | 15.12-20.8 | 20.81-25.45 | ≥25.46 |
| 11.25 | ≤13.04 | 13.05-15.16 | 15.17-20.88 | 20.89-25.56 | ≥25.57 |
| 11.33 | ≤13.08 | 13.09-15.2 | 15.21-20.95 | 20.96-25.66 | ≥25.67 |
| 11.42 | ≤13.11 | 13.12-15.25 | 15.26-21.03 | 21.04-25.76 | ≥25.77 |
| 11.5 | ≤13.15 | 13.16-15.3 | 15.31-21.11 | 21.12-25.86 | ≥25.87 |
| 11.58 | ≤13.18 | 13.19-15.35 | 15.36-21.19 | 21.2-25.97 | ≥25.98 |
| 11.67 | ≤13.22 | 13.23-15.39 | 15.4-21.26 | 21.27-26.07 | ≥26.08 |
| 11.75 | ≤13.26 | 13.27-15.44 | 15.45-21.34 | 21.35-26.17 | ≥26.18 |
| 11.83 | ≤13.3 | 13.31-15.49 | 15.5-21.42 | 21.43-26.27 | ≥26.28 |
| 11.92 | ≤13.34 | 13.35-15.54 | 15.55-21.5 | 21.51-26.37 | ≥26.38 |
| 12 | ≤13.38 | 13.39-15.59 | 15.6-21.58 | 21.59-26.46 | ≥26.47 |
| 12.08 | ≤13.42 | 13.43-15.65 | 15.66-21.65 | 21.66-26.56 | ≥26.57 |
| 12.17 | ≤13.47 | 13.48-15.7 | 15.71-21.73 | 21.74-26.66 | ≥26.67 |
| 12.25 | ≤13.51 | 13.52-15.75 | 15.76-21.81 | 21.82-26.75 | ≥26.76 |
| 12.33 | ≤13.55 | 13.56-15.8 | 15.81-21.89 | 21.9-26.85 | ≥26.86 |
| 12.42 | ≤13.6 | 13.61-15.86 | 15.87-21.96 | 21.97-26.94 | ≥26.95 |
| 12.5 | ≤13.64 | 13.65-15.91 | 15.92-22.04 | 22.05-27.04 | ≥27.05 |
| 12.58 | ≤13.69 | 13.7-15.96 | 15.97-22.11 | 22.12-27.13 | ≥27.14 |
| 12.67 | ≤13.73 | 13.74-16.02 | 16.03-22.19 | 22.2-27.21 | ≥27.22 |
| 12.75 | ≤13.78 | 13.79-16.07 | 16.08-22.26 | 22.27-27.3 | ≥27.31 |
| 12.83 | ≤13.82 | 13.83-16.13 | 16.14-22.34 | 22.35-27.39 | ≥27.4 |
| 12.92 | ≤13.87 | 13.88-16.18 | 16.19-22.41 | 22.42-27.48 | ≥27.49 |
| 13 | ≤13.92 | 13.93-16.23 | 16.24-22.48 | 22.49-27.56 | ≥27.57 |
| 13.08 | ≤13.96 | 13.97-16.29 | 16.3-22.55 | 22.56-27.64 | ≥27.65 |
| 13.17 | ≤14.01 | 14.02-16.34 | 16.35-22.62 | 22.63-27.72 | ≥27.73 |
| 13.25 | ≤14.06 | 14.07-16.4 | 16.41-22.69 | 22.7-27.8 | ≥27.81 |
| 13.33 | ≤14.1 | 14.11-16.45 | 16.46-22.76 | 22.77-27.87 | ≥27.88 |
| 13.42 | ≤14.15 | 14.16-16.5 | 16.51-22.83 | 22.84-27.95 | ≥27.96 |
| 13.5 | ≤14.2 | 14.21-16.55 | 16.56-22.89 | 22.9-28.02 | ≥28.03 |
| 13.58 | ≤14.24 | 14.25-16.61 | 16.62-22.96 | 22.97-28.09 | ≥28.1 |
| 13.67 | ≤14.29 | 14.3-16.66 | 16.67-23.02 | 23.03-28.15 | ≥28.16 |
| 13.75 | ≤14.34 | 14.35-16.71 | 16.72-23.08 | 23.09-28.22 | ≥28.23 |
| 13.83 | ≤14.38 | 14.39-16.76 | 16.77-23.14 | 23.15-28.28 | ≥28.29 |
| 13.92 | ≤14.43 | 14.44-16.81 | 16.82-23.2 | 23.21-28.35 | ≥28.36 |
| 14 | ≤14.47 | 14.48-16.86 | 16.87-23.26 | 23.27-28.41 | ≥28.42 |
| 14.08 | ≤14.52 | 14.53-16.91 | 16.92-23.32 | 23.33-28.47 | ≥28.48 |
| 14.17 | ≤14.57 | 14.58-16.96 | 16.97-23.38 | 23.39-28.52 | ≥28.53 |
| 14.25 | ≤14.61 | 14.62-17.01 | 17.02-23.43 | 23.44-28.58 | ≥28.59 |
| 14.33 | ≤14.65 | 14.66-17.06 | 17.07-23.49 | 23.5-28.63 | ≥28.64 |
| 14.42 | ≤14.7 | 14.71-17.11 | 17.12-23.54 | 23.55-28.68 | ≥28.69 |
| 14.5 | ≤14.74 | 14.75-17.16 | 17.17-23.59 | 23.6-28.73 | ≥28.74 |
| 14.58 | ≤14.79 | 14.8-17.2 | 17.21-23.64 | 23.65-28.78 | ≥28.79 |
| 14.67 | ≤14.83 | 14.84-17.25 | 17.26-23.69 | 23.7-28.83 | ≥28.84 |
| 14.75 | ≤14.87 | 14.88-17.3 | 17.31-23.74 | 23.75-28.87 | ≥28.88 |
| 14.83 | ≤14.92 | 14.93-17.34 | 17.35-23.79 | 23.8-28.91 | ≥28.92 |
| 14.92 | ≤14.96 | 14.97-17.39 | 17.4-23.83 | 23.84-28.96 | ≥28.97 |
| 15 | ≤15 | 15.01-17.43 | 17.44-23.88 | 23.89-29 | ≥29.01 |
| 15.08 | ≤15.04 | 15.05-17.47 | 17.48-23.92 | 23.93-29.04 | ≥29.05 |
| 15.17 | ≤15.08 | 15.09-17.51 | 17.52-23.96 | 23.97-29.07 | ≥29.08 |
| 15.25 | ≤15.12 | 15.13-17.56 | 17.57-24 | 24.01-29.11 | ≥29.12 |
| 15.33 | ≤15.16 | 15.17-17.6 | 17.61-24.04 | 24.05-29.14 | ≥29.15 |
| 15.42 | ≤15.2 | 15.21-17.64 | 17.65-24.08 | 24.09-29.18 | ≥29.19 |
| 15.5 | ≤15.24 | 15.25-17.68 | 17.69-24.12 | 24.13-29.21 | ≥29.22 |
| 15.58 | ≤15.27 | 15.28-17.72 | 17.73-24.16 | 24.17-29.24 | ≥29.25 |
| 15.67 | ≤15.31 | 15.32-17.75 | 17.76-24.2 | 24.21-29.28 | ≥29.29 |
| 15.75 | ≤15.34 | 15.35-17.79 | 17.8-24.23 | 24.24-29.3 | ≥29.31 |
| 15.83 | ≤15.38 | 15.39-17.82 | 17.83-24.27 | 24.28-29.33 | ≥29.34 |
| 15.92 | ≤15.41 | 15.42-17.86 | 17.87-24.3 | 24.31-29.36 | ≥29.37 |
| 16 | ≤15.45 | 15.46-17.9 | 17.91-24.33 | 24.34-29.39 | ≥29.4 |
| 16.08 | ≤15.48 | 15.49-17.93 | 17.94-24.37 | 24.38-29.41 | ≥29.42 |
| 16.17 | ≤15.51 | 15.52-17.96 | 17.97-24.4 | 24.41-29.44 | ≥29.45 |
| 16.25 | ≤15.54 | 15.55-17.99 | 18-24.43 | 24.44-29.47 | ≥29.48 |
| 16.33 | ≤15.57 | 15.58-18.02 | 18.03-24.46 | 24.47-29.49 | ≥29.5 |
| 16.42 | ≤15.6 | 15.61-18.06 | 18.07-24.49 | 24.5-29.52 | ≥29.53 |
| 16.5 | ≤15.63 | 15.64-18.08 | 18.09-24.52 | 24.53-29.54 | ≥29.55 |
| 16.58 | ≤15.65 | 15.66-18.11 | 18.12-24.55 | 24.56-29.57 | ≥29.58 |
| 16.67 | ≤15.68 | 15.69-18.14 | 18.15-24.58 | 24.59-29.59 | ≥29.6 |
| 16.75 | ≤15.7 | 15.71-18.17 | 18.18-24.6 | 24.61-29.62 | ≥29.63 |
| 16.83 | ≤15.73 | 15.74-18.19 | 18.2-24.63 | 24.64-29.64 | ≥29.65 |
| 16.92 | ≤15.75 | 15.76-18.22 | 18.23-24.66 | 24.67-29.67 | ≥29.68 |
| 17 | ≤15.78 | 15.79-18.24 | 18.25-24.69 | 24.7-29.69 | ≥29.7 |
| 17.08 | ≤15.8 | 15.81-18.27 | 18.28-24.71 | 24.72-29.72 | ≥29.73 |
| 17.17 | ≤15.82 | 15.83-18.29 | 18.3-24.74 | 24.75-29.74 | ≥29.75 |
| 17.25 | ≤15.84 | 15.85-18.31 | 18.32-24.76 | 24.77-29.76 | ≥29.77 |
| 17.33 | ≤15.86 | 15.87-18.34 | 18.35-24.79 | 24.8-29.79 | ≥29.8 |
| 17.42 | ≤15.88 | 15.89-18.36 | 18.37-24.81 | 24.82-29.81 | ≥29.82 |
| 17.5 | ≤15.9 | 15.91-18.38 | 18.39-24.84 | 24.85-29.84 | ≥29.85 |
| 17.58 | ≤15.91 | 15.92-18.4 | 18.41-24.87 | 24.88-29.86 | ≥29.87 |
| 17.67 | ≤15.93 | 15.94-18.42 | 18.43-24.89 | 24.9-29.89 | ≥29.9 |
| 17.75 | ≤15.95 | 15.96-18.44 | 18.45-24.92 | 24.93-29.91 | ≥29.92 |
| 17.83 | ≤15.97 | 15.98-18.46 | 18.47-24.94 | 24.95-29.94 | ≥29.95 |
| 17.92 | ≤15.98 | 15.99-18.48 | 18.49-24.97 | 24.98-29.97 | ≥29.98 |
| 18 | ≤16 | 16.01-18.5 | 18.51-24.99 | 25-29.99 | ≥30 |
| 18.08 | ≤16 | 16.01-18.5 | 18.51-24.99 | 25-29.99 | ≥30 |
| 18.17 | ≤16 | 16.01-18.5 | 18.51-24.99 | 25-29.99 | ≥30 |
| 18.25 | ≤16 | 16.01-18.5 | 18.51-24.99 | 25-29.99 | ≥30 |
| 18.33 | ≤16 | 16.01-18.5 | 18.51-24.99 | 25-29.99 | ≥30 |
| 18.42 | ≤16 | 16.01-18.5 | 18.51-24.99 | 25-29.99 | ≥30 |
| 18.5 | ≤16 | 16.01-18.5 | 18.51-24.99 | 25-29.99 | ≥30 |
| 18.58 | ≤16 | 16.01-18.5 | 18.51-24.99 | 25-29.99 | ≥30 |
| 18.67 | ≤16 | 16.01-18.5 | 18.51-24.99 | 25-29.99 | ≥30 |
| 18.75 | ≤16 | 16.01-18.5 | 18.51-24.99 | 25-29.99 | ≥30 |
| 18.83 | ≤16 | 16.01-18.5 | 18.51-24.99 | 25-29.99 | ≥30 |
| 18.92 | ≤16 | 16.01-18.5 | 18.51-24.99 | 25-29.99 | ≥30 |
| 19 | ≤16 | 16.01-18.5 | 18.51-24.99 | 25-29.99 | ≥30 |
| **BOYS** | | | | | |
| 5.08 | ≤12.78 | 12.79-14.24 | 14.25-17.38 | 17.39-19.28 | ≥19.29 |
| 5.17 | ≤12.75 | 12.76-14.22 | 14.23-17.39 | 17.4-19.31 | ≥19.32 |
| 5.25 | ≤12.73 | 12.74-14.2 | 14.21-17.39 | 17.4-19.34 | ≥19.35 |
| 5.33 | ≤12.71 | 12.72-14.18 | 14.19-17.4 | 17.41-19.37 | ≥19.38 |
| 5.42 | ≤12.69 | 12.7-14.17 | 14.18-17.4 | 17.41-19.41 | ≥19.42 |
| 5.5 | ≤12.66 | 12.67-14.15 | 14.16-17.41 | 17.42-19.45 | ≥19.46 |
| 5.58 | ≤12.64 | 12.65-14.13 | 14.14-17.43 | 17.44-19.49 | ≥19.5 |
| 5.67 | ≤12.62 | 12.63-14.11 | 14.12-17.44 | 17.45-19.54 | ≥19.55 |
| 5.75 | ≤12.6 | 12.61-14.1 | 14.11-17.45 | 17.46-19.58 | ≥19.59 |
| 5.83 | ≤12.58 | 12.59-14.08 | 14.09-17.47 | 17.48-19.64 | ≥19.65 |
| 5.92 | ≤12.56 | 12.57-14.07 | 14.08-17.49 | 17.5-19.69 | ≥19.7 |
| 6 | ≤12.54 | 12.55-14.06 | 14.07-17.51 | 17.52-19.75 | ≥19.76 |
| 6.08 | ≤12.52 | 12.53-14.04 | 14.05-17.53 | 17.54-19.81 | ≥19.82 |
| 6.17 | ≤12.5 | 12.51-14.03 | 14.04-17.55 | 17.56-19.87 | ≥19.88 |
| 6.25 | ≤12.48 | 12.49-14.02 | 14.03-17.58 | 17.59-19.93 | ≥19.94 |
| 6.33 | ≤12.47 | 12.48-14.01 | 14.02-17.61 | 17.62-20 | ≥20.01 |
| 6.42 | ≤12.45 | 12.46-14.01 | 14.02-17.63 | 17.64-20.07 | ≥20.08 |
| 6.5 | ≤12.44 | 12.45-14 | 14.01-17.66 | 17.67-20.14 | ≥20.15 |
| 6.58 | ≤12.43 | 12.44-14 | 14.01-17.69 | 17.7-20.21 | ≥20.22 |
| 6.67 | ≤12.42 | 12.43-13.99 | 14-17.72 | 17.73-20.28 | ≥20.29 |
| 6.75 | ≤12.41 | 12.42-13.99 | 14-17.76 | 17.77-20.35 | ≥20.36 |
| 6.83 | ≤12.4 | 12.41-13.99 | 14-17.79 | 17.8-20.43 | ≥20.44 |
| 6.92 | ≤12.39 | 12.4-13.99 | 14-17.83 | 17.84-20.5 | ≥20.51 |
| 7 | ≤12.39 | 12.4-14 | 14.01-17.87 | 17.88-20.58 | ≥20.59 |
| 7.08 | ≤12.39 | 12.4-14 | 14.01-17.9 | 17.91-20.65 | ≥20.66 |
| 7.17 | ≤12.39 | 12.4-14.01 | 14.02-17.94 | 17.95-20.73 | ≥20.74 |
| 7.25 | ≤12.39 | 12.4-14.02 | 14.03-17.98 | 17.99-20.81 | ≥20.82 |
| 7.33 | ≤12.39 | 12.4-14.02 | 14.03-18.03 | 18.04-20.89 | ≥20.9 |
| 7.42 | ≤12.39 | 12.4-14.04 | 14.05-18.07 | 18.08-20.97 | ≥20.98 |
| 7.5 | ≤12.39 | 12.4-14.05 | 14.06-18.11 | 18.12-21.05 | ≥21.06 |
| 7.58 | ≤12.4 | 12.41-14.06 | 14.07-18.16 | 18.17-21.13 | ≥21.14 |
| 7.67 | ≤12.4 | 12.41-14.07 | 14.08-18.2 | 18.21-21.21 | ≥21.22 |
| 7.75 | ≤12.41 | 12.42-14.09 | 14.1-18.25 | 18.26-21.29 | ≥21.3 |
| 7.83 | ≤12.41 | 12.42-14.1 | 14.11-18.3 | 18.31-21.38 | ≥21.39 |
| 7.92 | ≤12.42 | 12.43-14.12 | 14.13-18.35 | 18.36-21.46 | ≥21.47 |
| 8 | ≤12.43 | 12.44-14.13 | 14.14-18.4 | 18.41-21.55 | ≥21.56 |
| 8.08 | ≤12.44 | 12.45-14.15 | 14.16-18.45 | 18.46-21.64 | ≥21.65 |
| 8.17 | ≤12.44 | 12.45-14.17 | 14.18-18.5 | 18.51-21.73 | ≥21.74 |
| 8.25 | ≤12.45 | 12.46-14.18 | 14.19-18.55 | 18.56-21.82 | ≥21.83 |
| 8.33 | ≤12.46 | 12.47-14.2 | 14.21-18.61 | 18.62-21.91 | ≥21.92 |
| 8.42 | ≤12.47 | 12.48-14.22 | 14.23-18.66 | 18.67-22.01 | ≥22.02 |
| 8.5 | ≤12.48 | 12.49-14.24 | 14.25-18.72 | 18.73-22.1 | ≥22.11 |
| 8.58 | ≤12.49 | 12.5-14.26 | 14.27-18.77 | 18.78-22.2 | ≥22.21 |
| 8.67 | ≤12.5 | 12.51-14.28 | 14.29-18.83 | 18.84-22.3 | ≥22.31 |
| 8.75 | ≤12.51 | 12.52-14.3 | 14.31-18.89 | 18.9-22.4 | ≥22.41 |
| 8.83 | ≤12.52 | 12.53-14.32 | 14.33-18.94 | 18.95-22.5 | ≥22.51 |
| 8.92 | ≤12.53 | 12.54-14.34 | 14.35-19 | 19.01-22.6 | ≥22.61 |
| 9 | ≤12.54 | 12.55-14.36 | 14.37-19.06 | 19.07-22.7 | ≥22.71 |
| 9.08 | ≤12.55 | 12.56-14.38 | 14.39-19.12 | 19.13-22.81 | ≥22.82 |
| 9.17 | ≤12.56 | 12.57-14.4 | 14.41-19.18 | 19.19-22.91 | ≥22.92 |
| 9.25 | ≤12.58 | 12.59-14.42 | 14.43-19.24 | 19.25-23.02 | ≥23.03 |
| 9.33 | ≤12.59 | 12.6-14.44 | 14.45-19.3 | 19.31-23.12 | ≥23.13 |
| 9.42 | ≤12.6 | 12.61-14.47 | 14.48-19.36 | 19.37-23.23 | ≥23.24 |
| 9.5 | ≤12.61 | 12.62-14.49 | 14.5-19.42 | 19.43-23.33 | ≥23.34 |
| 9.58 | ≤12.63 | 12.64-14.51 | 14.52-19.48 | 19.49-23.44 | ≥23.45 |
| 9.67 | ≤12.64 | 12.65-14.53 | 14.54-19.54 | 19.55-23.54 | ≥23.55 |
| 9.75 | ≤12.65 | 12.66-14.56 | 14.57-19.6 | 19.61-23.65 | ≥23.66 |
| 9.83 | ≤12.67 | 12.68-14.58 | 14.59-19.66 | 19.67-23.75 | ≥23.76 |
| 9.92 | ≤12.68 | 12.69-14.61 | 14.62-19.73 | 19.74-23.85 | ≥23.86 |
| 10 | ≤12.7 | 12.71-14.63 | 14.64-19.79 | 19.8-23.95 | ≥23.96 |
| 10.08 | ≤12.71 | 12.72-14.66 | 14.67-19.85 | 19.86-24.05 | ≥24.06 |
| 10.17 | ≤12.73 | 12.74-14.68 | 14.69-19.91 | 19.92-24.15 | ≥24.16 |
| 10.25 | ≤12.74 | 12.75-14.71 | 14.72-19.96 | 19.97-24.24 | ≥24.25 |
| 10.33 | ≤12.76 | 12.77-14.73 | 14.74-20.03 | 20.04-24.34 | ≥24.35 |
| 10.42 | ≤12.78 | 12.79-14.76 | 14.77-20.08 | 20.09-24.43 | ≥24.44 |
| 10.5 | ≤12.8 | 12.81-14.79 | 14.8-20.14 | 20.15-24.53 | ≥24.54 |
| 10.58 | ≤12.81 | 12.82-14.82 | 14.83-20.2 | 20.21-24.62 | ≥24.63 |
| 10.67 | ≤12.83 | 12.84-14.84 | 14.85-20.26 | 20.27-24.71 | ≥24.72 |
| 10.75 | ≤12.85 | 12.86-14.87 | 14.88-20.32 | 20.33-24.8 | ≥24.81 |
| 10.83 | ≤12.87 | 12.88-14.9 | 14.91-20.38 | 20.39-24.89 | ≥24.9 |
| 10.92 | ≤12.89 | 12.9-14.93 | 14.94-20.44 | 20.45-24.97 | ≥24.98 |
| 11 | ≤12.91 | 12.92-14.96 | 14.97-20.5 | 20.51-25.06 | ≥25.07 |
| 11.08 | ≤12.94 | 12.95-14.99 | 15-20.55 | 20.56-25.14 | ≥25.15 |
| 11.17 | ≤12.96 | 12.97-15.02 | 15.03-20.61 | 20.62-25.23 | ≥25.24 |
| 11.25 | ≤12.98 | 12.99-15.05 | 15.06-20.67 | 20.68-25.31 | ≥25.32 |
| 11.33 | ≤13 | 13.01-15.08 | 15.09-20.73 | 20.74-25.39 | ≥25.4 |
| 11.42 | ≤13.03 | 13.04-15.12 | 15.13-20.78 | 20.79-25.47 | ≥25.48 |
| 11.5 | ≤13.05 | 13.06-15.15 | 15.16-20.84 | 20.85-25.55 | ≥25.56 |
| 11.58 | ≤13.08 | 13.09-15.18 | 15.19-20.9 | 20.91-25.63 | ≥25.64 |
| 11.67 | ≤13.1 | 13.11-15.22 | 15.23-20.96 | 20.97-25.71 | ≥25.72 |
| 11.75 | ≤13.13 | 13.14-15.25 | 15.26-21.02 | 21.03-25.78 | ≥25.79 |
| 11.83 | ≤13.16 | 13.17-15.29 | 15.3-21.07 | 21.08-25.86 | ≥25.87 |
| 11.92 | ≤13.19 | 13.2-15.32 | 15.33-21.13 | 21.14-25.93 | ≥25.94 |
| 12 | ≤13.21 | 13.22-15.36 | 15.37-21.19 | 21.2-26.01 | ≥26.02 |
| 12.08 | ≤13.24 | 13.25-15.4 | 15.41-21.24 | 21.25-26.08 | ≥26.09 |
| 12.17 | ≤13.28 | 13.29-15.44 | 15.45-21.3 | 21.31-26.16 | ≥26.17 |
| 12.25 | ≤13.31 | 13.32-15.47 | 15.48-21.36 | 21.37-26.23 | ≥26.24 |
| 12.33 | ≤13.34 | 13.35-15.51 | 15.52-21.42 | 21.43-26.3 | ≥26.31 |
| 12.42 | ≤13.37 | 13.38-15.55 | 15.56-21.48 | 21.49-26.37 | ≥26.38 |
| 12.5 | ≤13.4 | 13.41-15.59 | 15.6-21.53 | 21.54-26.44 | ≥26.45 |
| 12.58 | ≤13.44 | 13.45-15.63 | 15.64-21.59 | 21.6-26.51 | ≥26.52 |
| 12.67 | ≤13.47 | 13.48-15.67 | 15.68-21.65 | 21.66-26.58 | ≥26.59 |
| 12.75 | ≤13.5 | 13.51-15.71 | 15.72-21.71 | 21.72-26.65 | ≥26.66 |
| 12.83 | ≤13.54 | 13.55-15.75 | 15.76-21.77 | 21.78-26.72 | ≥26.73 |
| 12.92 | ≤13.58 | 13.59-15.8 | 15.81-21.82 | 21.83-26.79 | ≥26.8 |
| 13 | ≤13.61 | 13.62-15.84 | 15.85-21.88 | 21.89-26.86 | ≥26.87 |
| 13.08 | ≤13.65 | 13.66-15.88 | 15.89-21.94 | 21.95-26.93 | ≥26.94 |
| 13.17 | ≤13.69 | 13.7-15.93 | 15.94-22 | 22.01-26.99 | ≥27 |
| 13.25 | ≤13.73 | 13.74-15.97 | 15.98-22.06 | 22.07-27.06 | ≥27.07 |
| 13.33 | ≤13.76 | 13.77-16.02 | 16.03-22.12 | 22.13-27.13 | ≥27.14 |
| 13.42 | ≤13.8 | 13.81-16.06 | 16.07-22.18 | 22.19-27.19 | ≥27.2 |
| 13.5 | ≤13.84 | 13.85-16.11 | 16.12-22.23 | 22.24-27.25 | ≥27.26 |
| 13.58 | ≤13.88 | 13.89-16.16 | 16.17-22.29 | 22.3-27.32 | ≥27.33 |
| 13.67 | ≤13.93 | 13.94-16.2 | 16.21-22.35 | 22.36-27.38 | ≥27.39 |
| 13.75 | ≤13.97 | 13.98-16.25 | 16.26-22.41 | 22.42-27.45 | ≥27.46 |
| 13.83 | ≤14.01 | 14.02-16.3 | 16.31-22.47 | 22.48-27.51 | ≥27.52 |
| 13.92 | ≤14.05 | 14.06-16.35 | 16.36-22.53 | 22.54-27.57 | ≥27.58 |
| 14 | ≤14.09 | 14.1-16.39 | 16.4-22.59 | 22.6-27.63 | ≥27.64 |
| 14.08 | ≤14.14 | 14.15-16.44 | 16.45-22.65 | 22.66-27.69 | ≥27.7 |
| 14.17 | ≤14.18 | 14.19-16.49 | 16.5-22.71 | 22.72-27.75 | ≥27.76 |
| 14.25 | ≤14.22 | 14.23-16.54 | 16.55-22.76 | 22.77-27.81 | ≥27.82 |
| 14.33 | ≤14.26 | 14.27-16.59 | 16.6-22.82 | 22.83-27.87 | ≥27.88 |
| 14.42 | ≤14.31 | 14.32-16.64 | 16.65-22.88 | 22.89-27.93 | ≥27.94 |
| 14.5 | ≤14.35 | 14.36-16.68 | 16.69-22.94 | 22.95-27.99 | ≥28 |
| 14.58 | ≤14.4 | 14.41-16.73 | 16.74-22.99 | 23-28.04 | ≥28.05 |
| 14.67 | ≤14.44 | 14.45-16.78 | 16.79-23.05 | 23.06-28.1 | ≥28.11 |
| 14.75 | ≤14.48 | 14.49-16.83 | 16.84-23.11 | 23.12-28.15 | ≥28.16 |
| 14.83 | ≤14.53 | 14.54-16.88 | 16.89-23.16 | 23.17-28.21 | ≥28.22 |
| 14.92 | ≤14.57 | 14.58-16.93 | 16.94-23.22 | 23.23-28.26 | ≥28.27 |
| 15 | ≤14.61 | 14.62-16.98 | 16.99-23.27 | 23.28-28.31 | ≥28.32 |
| 15.08 | ≤14.66 | 14.67-17.02 | 17.03-23.32 | 23.33-28.36 | ≥28.37 |
| 15.17 | ≤14.7 | 14.71-17.07 | 17.08-23.38 | 23.39-28.41 | ≥28.42 |
| 15.25 | ≤14.74 | 14.75-17.12 | 17.13-23.43 | 23.44-28.46 | ≥28.47 |
| 15.33 | ≤14.78 | 14.79-17.16 | 17.17-23.48 | 23.49-28.51 | ≥28.52 |
| 15.42 | ≤14.83 | 14.84-17.21 | 17.22-23.53 | 23.54-28.55 | ≥28.56 |
| 15.5 | ≤14.87 | 14.88-17.26 | 17.27-23.58 | 23.59-28.6 | ≥28.61 |
| 15.58 | ≤14.91 | 14.92-17.3 | 17.31-23.63 | 23.64-28.65 | ≥28.66 |
| 15.67 | ≤14.95 | 14.96-17.35 | 17.36-23.68 | 23.69-28.69 | ≥28.7 |
| 15.75 | ≤15 | 15.01-17.4 | 17.41-23.73 | 23.74-28.74 | ≥28.75 |
| 15.83 | ≤15.04 | 15.05-17.44 | 17.45-23.78 | 23.79-28.79 | ≥28.8 |
| 15.92 | ≤15.08 | 15.09-17.49 | 17.5-23.83 | 23.84-28.83 | ≥28.84 |
| 16 | ≤15.12 | 15.13-17.53 | 17.54-23.88 | 23.89-28.88 | ≥28.89 |
| 16.08 | ≤15.16 | 15.17-17.57 | 17.58-23.93 | 23.94-28.92 | ≥28.93 |
| 16.17 | ≤15.2 | 15.21-17.62 | 17.63-23.98 | 23.99-28.96 | ≥28.97 |
| 16.25 | ≤15.24 | 15.25-17.66 | 17.67-24.03 | 24.04-29.01 | ≥29.02 |
| 16.33 | ≤15.28 | 15.29-17.71 | 17.72-24.07 | 24.08-29.05 | ≥29.06 |
| 16.42 | ≤15.32 | 15.33-17.75 | 17.76-24.12 | 24.13-29.1 | ≥29.11 |
| 16.5 | ≤15.36 | 15.37-17.79 | 17.8-24.17 | 24.18-29.14 | ≥29.15 |
| 16.58 | ≤15.4 | 15.41-17.83 | 17.84-24.21 | 24.22-29.19 | ≥29.2 |
| 16.67 | ≤15.44 | 15.45-17.88 | 17.89-24.26 | 24.27-29.23 | ≥29.24 |
| 16.75 | ≤15.47 | 15.48-17.92 | 17.93-24.31 | 24.32-29.28 | ≥29.29 |
| 16.83 | ≤15.51 | 15.52-17.96 | 17.97-24.36 | 24.37-29.33 | ≥29.34 |
| 16.92 | ≤15.55 | 15.56-18 | 18.01-24.4 | 24.41-29.37 | ≥29.38 |
| 17 | ≤15.59 | 15.6-18.04 | 18.05-24.45 | 24.46-29.42 | ≥29.43 |
| 17.08 | ≤15.62 | 15.63-18.08 | 18.09-24.49 | 24.5-29.47 | ≥29.48 |
| 17.17 | ≤15.66 | 15.67-18.12 | 18.13-24.54 | 24.55-29.51 | ≥29.52 |
| 17.25 | ≤15.69 | 15.7-18.16 | 18.17-24.59 | 24.6-29.56 | ≥29.57 |
| 17.33 | ≤15.73 | 15.74-18.2 | 18.21-24.63 | 24.64-29.61 | ≥29.62 |
| 17.42 | ≤15.76 | 15.77-18.24 | 18.25-24.68 | 24.69-29.66 | ≥29.67 |
| 17.5 | ≤15.8 | 15.81-18.28 | 18.29-24.72 | 24.73-29.7 | ≥29.71 |
| 17.58 | ≤15.83 | 15.84-18.31 | 18.32-24.77 | 24.78-29.75 | ≥29.76 |
| 17.67 | ≤15.87 | 15.88-18.35 | 18.36-24.81 | 24.82-29.8 | ≥29.81 |
| 17.75 | ≤15.9 | 15.91-18.39 | 18.4-24.86 | 24.87-29.85 | ≥29.86 |
| 17.83 | ≤15.93 | 15.94-18.43 | 18.44-24.9 | 24.91-29.89 | ≥29.9 |
| 17.92 | ≤15.97 | 15.98-18.46 | 18.47-24.95 | 24.96-29.94 | ≥29.95 |
| 18 | ≤16 | 16.01-18.5 | 18.51-24.99 | 25-29.99 | ≥30 |
| 18.08 | ≤16 | 16.01-18.5 | 18.51-24.99 | 25-29.99 | ≥30 |
| 18.17 | ≤16 | 16.01-18.5 | 18.51-24.99 | 25-29.99 | ≥30 |
| 18.25 | ≤16 | 16.01-18.5 | 18.51-24.99 | 25-29.99 | ≥30 |
| 18.33 | ≤16 | 16.01-18.5 | 18.51-24.99 | 25-29.99 | ≥30 |
| 18.42 | ≤16 | 16.01-18.5 | 18.51-24.99 | 25-29.99 | ≥30 |
| 18.5 | ≤16 | 16.01-18.5 | 18.51-24.99 | 25-29.99 | ≥30 |
| 18.58 | ≤16 | 16.01-18.5 | 18.51-24.99 | 25-29.99 | ≥30 |
| 18.67 | ≤16 | 16.01-18.5 | 18.51-24.99 | 25-29.99 | ≥30 |
| 18.75 | ≤16 | 16.01-18.5 | 18.51-24.99 | 25-29.99 | ≥30 |
| 18.83 | ≤16 | 16.01-18.5 | 18.51-24.99 | 25-29.99 | ≥30 |
| 18.92 | ≤16 | 16.01-18.5 | 18.51-24.99 | 25-29.99 | ≥30 |
| 19 | ≤16 | 16.01-18.5 | 18.51-24.99 | 25-29.99 | ≥30 |

**Table S7. FitBack health-related reference values for waist circumference (cm) by age group, stratified by sex (based on Jolliffe, C. J., & Janssen, I. (2007). Development of age-specific adolescent metabolic syndrome criteria that are linked to the adult treatment panel III and international diabetes federation criteria. Journal of the American College of Cardiology, 49(8), 891-898. doi:10.1016/j.jacc.2006.08.065)**

| **Age (Years)** | **Fit Zone** | **Improvement recommended** | **Improvement strongly recommended** |
| --- | --- | --- | --- |
| **GIRLS** | | | |
| 6 | ≤59.1 | 59.2-63.5 | ≥63.6 |
| 7 | ≤62.0 | 62.1-67.2 | ≥67.3 |
| 8 | ≤65.1 | 65.2-70.0 | ≥71.1 |
| 9 | ≤67.3 | 67.4-73.8 | ≥73.9 |
| 10 | ≤69.3 | 69.4-76.1 | ≥76.2 |
| 11 | ≤71.3 | 71.4-78.3 | ≥78.4 |
| 12 | ≤72.5 | 72.6-79.4 | ≥79.5 |
| 13 | ≤74.2 | 74.3-81.2 | ≥81.3 |
| 14 | ≤75.7 | 75.8-82.8 | ≥82.9 |
| 15 | ≤76.8 | 76.9-84.1 | ≥84.2 |
| 16 | ≤77.7 | 77.8-85.1 | ≥85.2 |
| 17 | ≤78.5 | 78.6-86.1 | ≥86.2 |
| 18 | ≤79.2 | 79.3-86.9 | ≥87.0 |
| 19 | ≤79.8 | 79.9-87.6 | ≥87.7 |
| **BOYS** | | | |
| 6 | ≤65.7 | 65.8-70.3 | ≥70.4 |
| 7 | ≤69.6 | 69.7-74.9 | ≥75 |
| 8 | ≤74 | 74.1-80.2 | ≥80.3 |
| 9 | ≤77.7 | 77.8-84.6 | ≥84.7 |
| 10 | ≤80.8 | 80.9-88.4 | ≥88.5 |
| 11 | ≤83.5 | 83.6-91.5 | ≥91.6 |
| 12 | ≤85.1 | 85.2-94.1 | ≥94.2 |
| 13 | ≤87 | 87.1-96.1 | ≥96.2 |
| 14 | ≤88.9 | 89-97.9 | ≥98 |
| 15 | ≤90.5 | 90.6-99.4 | ≥99.5 |
| 16 | ≤91.8 | 91.9-100.5 | ≥100.6 |
| 17 | ≤92.7 | 92.8-101.3 | ≥101.4 |
| 18 | ≤93.4 | 93.5-101.7 | ≥101.8 |
| 19 | ≤93.8 | 93.9-101.9 | ≥102 |

**Table S8. FitBack health-related reference values for 20m shuttle run test by age group, stratified by sex. Values shown are VO_2_max (ml/kg/min) which is calculated according to equation proposed by Nevill et al, 2020. (Based on Ruiz, J. R., Cavero-Redondo, I., Ortega, F. B., Welk, G. J., Andersen, L. B., & Martinez-Vizcaino, V. (2016). Cardiorespiratory fitness cut points to avoid cardiovascular disease risk in children and adolescents; what level of fitness should raise a red flag? A systematic review and meta-analysis. British Journal of Sports Medicine, 50(23), 1451-1458. doi:10.1136/bjsports-2015-095903).**

| **Age (Years)** | **Fit Zone** | **Improvement recommended** | **Improvement strongly recommended** |
| --- | --- | --- | --- |
| **GIRLS** | | | |
| 6 | ≥39.5 | 34.61-39.49 | ≤34.6 |
| 7 | ≥39.5 | 34.61-39.49 | ≤34.6 |
| 8 | ≥39.5 | 34.61-39.49 | ≤34.6 |
| 9 | ≥39.5 | 34.61-39.49 | ≤34.6 |
| 10 | ≥39.5 | 34.61-39.49 | ≤34.6 |
| 11 | ≥39.5 | 34.61-39.49 | ≤34.6 |
| 12 | ≥39.5 | 34.61-39.49 | ≤34.6 |
| 13 | ≥39.5 | 34.61-39.49 | ≤34.6 |
| 14 | ≥39.5 | 34.61-39.49 | ≤34.6 |
| 15 | ≥39.5 | 34.61-39.49 | ≤34.6 |
| 16 | ≥39.5 | 34.61-39.49 | ≤34.6 |
| 17 | ≥39.5 | 34.61-39.49 | ≤34.6 |
| 18 | ≥39.5 | 34.61-39.49 | ≤34.6 |
| 19 | ≥39.5 | 34.61-39.49 | ≤34.6 |
| **BOYS** | | | |
| 6 | ≥47.0 | 41.81-46.99 | ≤41.8 |
| 7 | ≥47.0 | 41.81-46.99 | ≤41.8 |
| 8 | ≥47.0 | 41.81-46.99 | ≤41.8 |
| 9 | ≥47.0 | 41.81-46.99 | ≤41.8 |
| 10 | ≥47.0 | 41.81-46.99 | ≤41.8 |
| 11 | ≥47.0 | 41.81-46.99 | ≤41.8 |
| 12 | ≥47.0 | 41.81-46.99 | ≤41.8 |
| 13 | ≥47.0 | 41.81-46.99 | ≤41.8 |
| 14 | ≥47.0 | 41.81-46.99 | ≤41.8 |
| 15 | ≥47.0 | 41.81-46.99 | ≤41.8 |
| 16 | ≥47.0 | 41.81-46.99 | ≤41.8 |
| 17 | ≥47.0 | 41.81-46.99 | ≤41.8 |
| 18 | ≥47.0 | 41.81-46.99 | ≤41.8 |
| 19 | ≥47.0 | 41.81-46.99 | ≤41.8 |

**Table S9. FitBack health-realted reference values for handgrip strength (kg) by age group, stratified by sex (based on Saint-Maurice, P. F., Laurson, K., Welk, G. J., Eisenmann, J., Gracia-Marco, L., Artero, E. G., . . . Janz, K. F. (2018). Grip strength cutpoints for youth based on a clinically relevant bone health outcome. Archives of Osteoporosis, 13(1) doi:10.1007/s11657-018-0502-0)**

| **Age (Years)** | **Fit Zone** | **Improvement recommended** | **Improvement strongly recommended** |
| --- | --- | --- | --- |
| **GIRLS** | | | |
| 6 | ≥8.9 | 7.41-8.89 | ≤7.4 |
| 7 | ≥10.2 | 8.51-10.19 | ≤8.5 |
| 8 | ≥12 | 9.91-11.99 | ≤9.9 |
| 9 | ≥14 | 11.61-13.99 | ≤11.6 |
| 10 | ≥16.5 | 13.61-16.49 | ≤13.6 |
| 11 | ≥19.4 | 16.01-19.39 | ≤16 |
| 12 | ≥22.1 | 18.21-22.09 | ≤18.2 |
| 13 | ≥24.2 | 19.91-24.19 | ≤19.9 |
| 14 | ≥25.7 | 21.21-25.69 | ≤21.2 |
| 15 | ≥26.8 | 22.11-26.79 | ≤22.1 |
| 16 | ≥27.6 | 22.71-27.59 | ≤22.7 |
| 17 | ≥28.3 | 23.31-28.29 | ≤23.3 |
| 18 | ≥28.7 | 23.71-28.69 | ≤23.7 |
| 19 | ≥29.4 | 24.51-29.39 | ≤24.5 |
| **BOYS** | | | |
| 6 | ≥10 | 7.81-9.99 | ≤7.8 |
| 7 | ≥11.2 | 8.91-11.19 | ≤8.9 |
| 8 | ≥13.1 | 10.41-13.09 | ≤10.4 |
| 9 | ≥15.2 | 12.01-15.19 | ≤12 |
| 10 | ≥17.3 | 13.81-17.29 | ≤13.8 |
| 11 | ≥19.9 | 15.81-19.89 | ≤15.8 |
| 12 | ≥23.1 | 18.41-23.09 | ≤18.4 |
| 13 | ≥27.4 | 21.91-27.39 | ≤21.9 |
| 14 | ≥32.2 | 25.71-32.19 | ≤25.7 |
| 15 | ≥36.3 | 29.01-36.29 | ≤29 |
| 16 | ≥39.3 | 31.51-39.29 | ≤31.5 |

**Table S10. FitBack health-related reference values for standing long jump (cm) by age group, stratified by sex (Based on Castro-Piñero, J., Laurson, K. R., Artero, E. G., Ortega, F. B., Labayen, I., Ruperez, A. I., . . . Ruiz, J. R. (2019). Muscle strength field-based tests to identify european adolescents at risk of metabolic syndrome: The HELENA study. Journal of Science and Medicine in Sport, 22(8), 929-934. doi:10.1016/j.jsams.2019.04.008)**

| **Age (Years)** | **Fit Zone** | **Improvement recommended** | **Improvement strongly recommended** |
| --- | --- | --- | --- |
| **GIRLS** | | | |
| 6 | ≥84.7 | 67.8-84.6 | ≤67.7 |
| 7 | ≥92.3 | 75-92.2 | ≤74.9 |
| 8 | ≥100.7 | 82.8-100.6 | ≤82.7 |
| 9 | ≥109.3 | 91.1-109.2 | ≤91 |
| 10 | ≥117.2 | 98.5-117.1 | ≤98.4 |
| 11 | ≥125.3 | 105.7-125.2 | ≤105.6 |
| 12 | ≥133.5 | 112.6-133.4 | ≤112.5 |
| 13 | ≥139.8 | 118.2-139.7 | ≤118.1 |
| 14 | ≥143.7 | 121.9-143.6 | ≤121.8 |
| 15 | ≥145.3 | 123.1-145.2 | ≤123 |
| 16 | ≥145.1 | 126.1-145 | ≤126 |
| 17 | ≥145.1 | 129.6-145 | ≤129.5 |
| 18 | ≥146.1 | 131-146 | ≤130.9 |
| 19 | ≥147.6 | 132.7-147.5 | ≤132.6 |
| **BOYS** | | | |
| 6 | ≥92.4 | 77.9-92.3 | ≤77.8 |
| 7 | ≥100.5 | 85.7-100.4 | ≤85.6 |
| 8 | ≥110.1 | 94.7-110 | ≤94.6 |
| 9 | ≥119.3 | 103.4-119.2 | ≤103.3 |
| 10 | ≥127.4 | 111-127.3 | ≤110.9 |
| 11 | ≥134.9 | 117.9-134.8 | ≤117.8 |
| 12 | ≥143.5 | 125.4-143.4 | ≤125.3 |
| 13 | ≥154.6 | 135.6-154.5 | ≤135.5 |
| 14 | ≥167.7 | 151.7-167.6 | ≤151.6 |
| 15 | ≥179.4 | 165.6-179.3 | ≤165.5 |
| 16 | ≥187.7 | 175.2-187.6 | ≤175.1 |
| 17 | ≥193 | 184.4-192.9 | ≤184.3 |
| 18 | ≥196.8 | 188.5-196.7 | ≤188.4 |
| 19 | ≥200 | 191.5-199.9 | ≤191.4 |

Boys

|  | NI-thin Fitnessgram | NI- obesity - Fitnessgram | NI-thin - Fitback | NI- obesity - Fitback |
| --- | --- | --- | --- | --- |
| 6 | 13.7 | 17.8 | 12.54 | 19.76 |
| 7 | 13.7 | 18.3 | 12.39 | 20.59 |
| 8 | 13.8 | 19 | 12.43 | 21.56 |
| 9 | 14 | 19.9 | 12.54 | 22.71 |
| 10 | 14.2 | 20.8 | 12.7 | 23.96 |
| 11 | 14.5 | 21.8 | 12.91 | 25.07 |
| 12 | 15 | 22.7 | 13.21 | 26.02 |
| 13 | 15.4 | 23.6 | 13.61 | 26.87 |
| 14 | 16 | 24.5 | 14.09 | 27.64 |
| 15 | 16.5 | 25.3 | 14.61 | 28.32 |
| 16 | 17.1 | 26 | 15.12 | 28.89 |
| 17 | 17.7 | 26.7 | 15.59 | 29.43 |
| 18 | 18.2 | 27.5 | 16 | 30 |
| 19 | 18.2 | 27.5 | 16 | 30 |

Girls

|  | NI-thin Fitnessgram | NI- obesity - Fitnessgram | NI-thin - Fitback | NI- obesity - Fitback |
| --- | --- | --- | --- | --- |
| 6 | 13.4 | 17.7 | 12.34 | 19.61 |
| 7 | 13.4 | 18.3 | 12.23 | 20.39 |
| 8 | 13.5 | 19.1 | 12.3 | 21.44 |
| 9 | 13.7 | 20 | 12.44 | 22.66 |
| 10 | 14 | 21 | 12.63 | 23.97 |
| 11 | 14.4 | 21.9 | 12.94 | 25.25 |
| 12 | 14.8 | 22.9 | 13.38 | 26.47 |
| 13 | 15.3 | 23.8 | 13.92 | 27.57 |
| 14 | 15.8 | 24.6 | 14.47 | 28.42 |
| 15 | 16.3 | 25.4 | 15 | 29.01 |
| 16 | 16.8 | 26.1 | 15.45 | 29.4 |
| 17 | 17.2 | 26.7 | 15.78 | 29.7 |
| 18 | 17.5 | 27.2 | 16 | 30 |
| 19 | 17.5 | 27.2 | 16 | 30 |

**Figure S2. Comparison of Fitback health-related criteria for Body mass index (solid lines) with Fitnessgram reference values (dashed lines; Welk GJ, Going SB, Morrow JR Jr, Meredith MD. Development of new criterion-referenced fitness standards in the FITNESSGRAM® program: rationale and conceptual overview. Am J Prev Med. 2011 Oct;41(4 Suppl 2):S63-7. doi: 10.1016/j.amepre.2011.07.012.)**

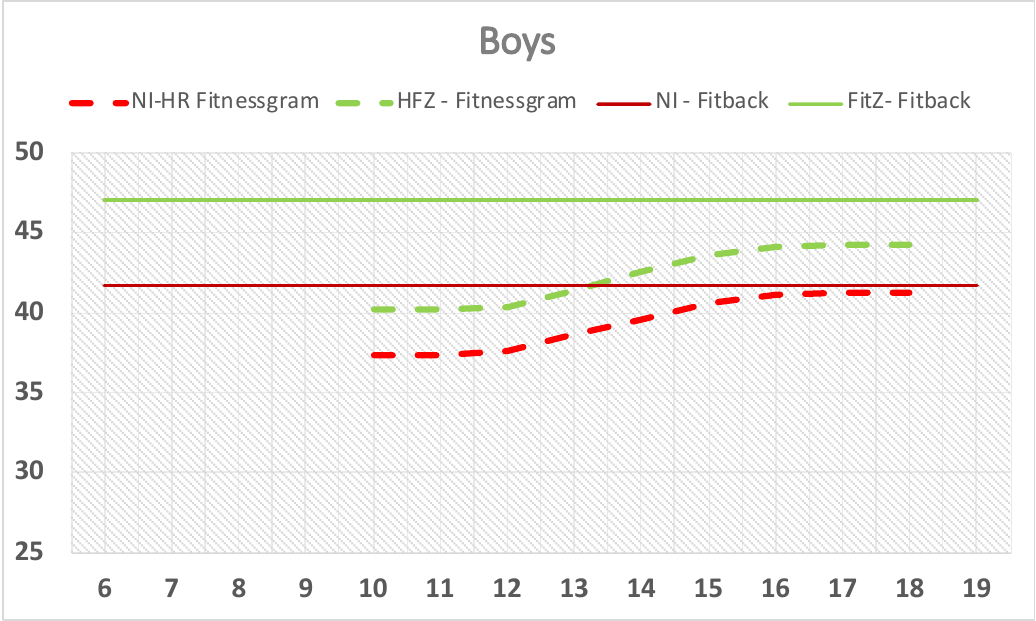

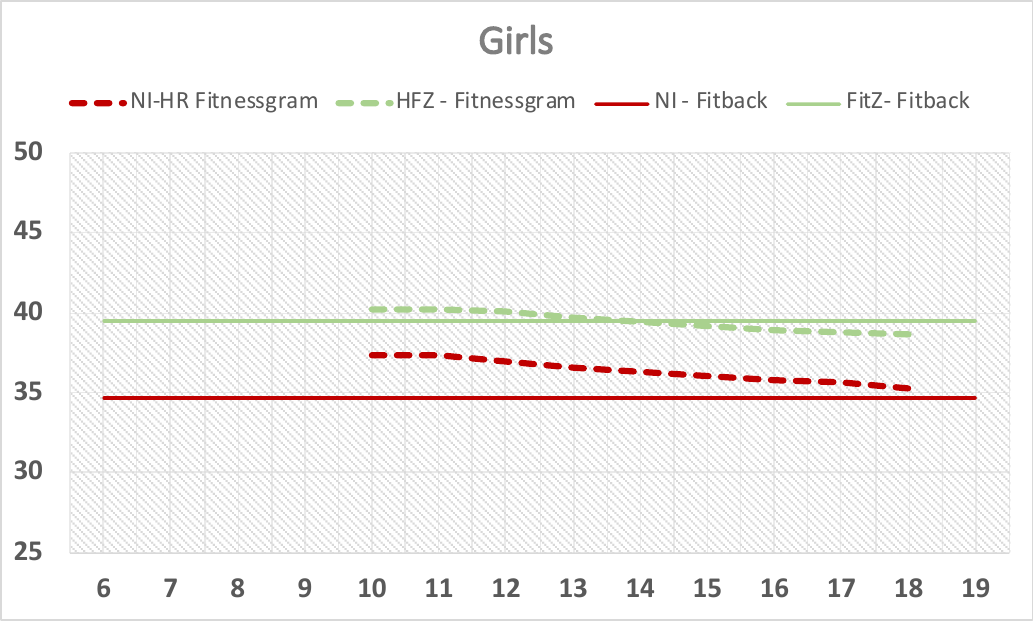

**Figure S3. Comparison of Fitback health-related criteria (solid lines) for cardiorespiratory fitness (operationalised through peak oxygen uptake in mlO_2_/kg/min) with Fitnessgram reference values (dashed lines; Welk GJ, Going SB, Morrow JR Jr, Meredith MD. Development of new criterion-referenced fitness standards in the FITNESSGRAM® program: rationale and conceptual overview. Am J Prev Med. 2011 Oct;41(4 Suppl 2):S63-7. doi: 10.1016/j.amepre.2011.07.01**
